# Development of a predictive algorithm for patient survival after traumatic injury using a five analyte blood panel

**DOI:** 10.1101/2024.04.22.24306188

**Authors:** Parinaz Fathi, Maria Karkanitsa, Adam Rupert, Aaron Lin, Jenna Darrah, F. Dennis Thomas, Jeffrey Lai, Kavita Babu, Mark Neavyn, Rosemary Kozar, Christopher Griggs, Kyle W. Cunningham, Carl I. Schulman, Marie Crandall, Irini Sereti, Emily Ricotta, Kaitlyn Sadtler

**Affiliations:** Section on Immunoengineering, Center for Biomedical Engineering and Technology Acceleration, National Institute of Biomedical Imaging and Bioengineering (NIBIB), National Institutes of Health (NIH), Bethesda, MD 20892; Unit for Nanoengineering and Microphysiologic Systems, NIBIB, NIH, Bethesda MD 20892; AIDS Monitoring Laboratory, Frederick National Laboratory for Cancer Research, Frederick MD; Dunlap and Associates, Inc., Cary, NC, 27511; Department of Emergency Medicine, University of Massachusetts Medical School, Worcester MA 01655; Shock Trauma Center, University of Maryland School of Medicine, Baltimore MD 21201; Department of Emergency Medicine, Atrium Health’s Carolinas Medical Center, Charlotte NC 28203; Division of Acute Care Surgery, Atrium Health’s Carolinas Medical Center, Charlotte NC 28203; University of Miami Miller School of Medicine, Miami FL 33136; Department of Surgery, University of Florida College of Medicine, Jacksonville FL 33209; Laboratory of Immunoregulation, Division of Intramural Research, National Institute of Allergy and Infectious Diseases (NIAID), NIH; Epidemiology and Data Management Unit, Laboratory of Clinical Immunology and Microbiology, NIAID, NIH, Bethesda, MD 20892; Preventative Medicine and Biostatistics, Uniformed Services University of the Health Sciences, Bethesda MD 20814

## Abstract

Severe trauma can induce systemic inflammation but also immunosuppression, which makes understanding the immune response of trauma patients critical for therapeutic development and treatment approaches. By evaluating the levels of 59 proteins in the plasma of 50 healthy volunteers and 1000 trauma patients across five trauma centers in the United States, we identified 6 novel changes in immune proteins after traumatic injury and further new variations by sex, age, trauma type, comorbidities, and developed a new equation for prediction of patient survival. Blood was collected at the time of arrival at Level 1 trauma centers and patients were stratified based on trauma level, tissues injured, and injury types. Trauma patients had significantly upregulated proteins associated with immune activation (IL-23, MIP-5), immunosuppression (IL-10) and pleiotropic cytokines (IL-29, IL-6). A high ratio of IL-29 to IL-10 was identified as a new predictor of survival in less severe patients with ROC area of 0.933. Combining machine learning with statistical modeling we developed an equation (“VIPER”) that could predict survival with ROC 0.966 in less severe patients and 0.8873 for all patients from a five analyte panel (IL-6, VEGF-A, IL-21, IL-29, and IL-10). Furthermore, we also identified three increased proteins (MIF, TRAIL, IL-29) and three decreased proteins (IL-7, TPO, IL-8) that were the most important in distinguishing a trauma blood profile. Biologic sex altered phenotype with IL-8 and MIF being lower in healthy women, but higher in female trauma patients when compared to male counterparts. This work identifies new responses to injury that may influence systemic immune dysfunction, serving as targets for therapeutics and immediate clinical benefit in identifying at-risk patients.

## INTRODUCTION

Traumatic injury leads to a cascade of immune activation to prevent infection and scavenge debris from damaged tissues, followed by a refractory immunosuppressive response due to increases in glucocorticoids and responses to the initial inflammatory activation. This natural immune suppression can increase susceptibility to downstream infections at the injury site as well as opportunistic infections such as respiratory viruses and bacterial pneumonia. The large number of variables – both intrinsic to proteins and cells of the immune response and extrinsic to the injury itself – complicates inferences about factors affecting patient outcomes and efforts to develop therapeutic agents to improve treatment. By assembling a large, diverse cohort of trauma patients, we can elucidate the immune phenotypes associated with traumatic injury and their role in and prediction of trauma recovery.

Recent technological advances have increased our understanding of the human immune response to traumatic injury.^1^ In addition to glucocorticoid release, a significant increase in interleukin-10 (IL-10) is associated with immunoregulation and immunosuppression. Increasing severity of wounds is associated with increasing systemic (peripheral blood) concentrations of IL-10.^2^ Other cytokines such as IL-4, IL-6, IL-8, and transforming growth factor-beta (TGFβ) have been implicated in systemic inflammatory response after trauma.^3^ Severe injury has been shown to correlate with downregulation of IL-9, IL-21, IL-22, IL-23, and IL-17E/IL-25 in blunt trauma patients, all of which are mediators of tissue repair.^4^

While tissue-based analyses often yield more insight into the immune function in response to specific injury types, peripheral blood, despite being limited to providing systemic rather than local information, is a more easily accessible analytic medium that can be collected longitudinally and less invasively. An ability to infer outcomes or inform therapeutic decision making from peripheral blood proteins can lead to potential diagnostic platforms for use in triage, allow more insight into the basic mechanistic responses to injury, and provide investigative targets to explore for downline therapeutic engineering.^5,6^ Here, we present a large, 1000 patient retrospective study on human systemic immune response to trauma, evaluating 59 plasma proteins in comparison to healthy controls, and stratified by demographics, mechanism of injury, body region, and tissues injured. These plasma proteins were selected to include proteins associated with inflammation and immunity whose levels were detectable with multiplexed assays based on preliminary analyses of a small subset of samples. Furthermore, we utilize machine learning to classify proteins that are important for predicting a trauma-associated inflammatory profile and identify a novel predictor of patient survival that can have immediate clinical impact. This work represents the largest study of plasma proteins in trauma patients and provides insight that can be used for patient diagnosis and prediction of patient outcomes, as well as a potential therapeutic pathway for further research.

## RESULTS

We analyzed 1000 plasma samples collected as a part of astudy focused on drug prevalence among trauma victims treated at selected Level 1 trauma centers^7–9^ and 50 samples from healthy individuals at the NIH. Trauma patient samples came from 5 centers located in Worcester MA, Baltimore MD, Charlotte NC, Jacksonville FL, and Miami FL. Of the trauma samples, 997 had complete clinical and laboratory data (**Supplementary Figure 1**). The median age of trauma patients was 39 years (**Figure 1a**, **Supplemental Table 1**). The majority of patients were male (71.8%) and White/European American (55.1%). Black/African Americans represented 36.9%, other races comprised 1.6%, and race was unknown for the remaining 6.2% of patients (**Figure 1b**). Many patients were seen in the emergency department and discharged (60.3%); 25% were sent to the operating room (OR), 12.3% to the intensive care unit (ICU), and 2.5% ultimately died from their injuries (**Fig. 1b**). Almost half (42%) of patients were admitted due to a motor vehicle crash (MVC) followed by falls (21.1%) and gunshot/shotgun wounds (GSW, 11.2%). Injury types included fractures (57.9%), lacerations (40.7%), abrasions (31.5%), and contusions (23.6%) with many patients reporting more than one injury type (**Figure 1c**). Additionally, we categorized injuries as “skin breaking” (avulsion, incision, laceration, open fracture, stab wound, puncture, gunshot wound, amputation, abrasion, and burns) or “internal” (hematoma, ecchymosis, contusion, seatbelt mark, swelling, fracture, deformity) to evaluate tissue-specific immune responses (**Figure 1c**). Additional details are in **Supplemental Figure 1**.

**Figure 1.**
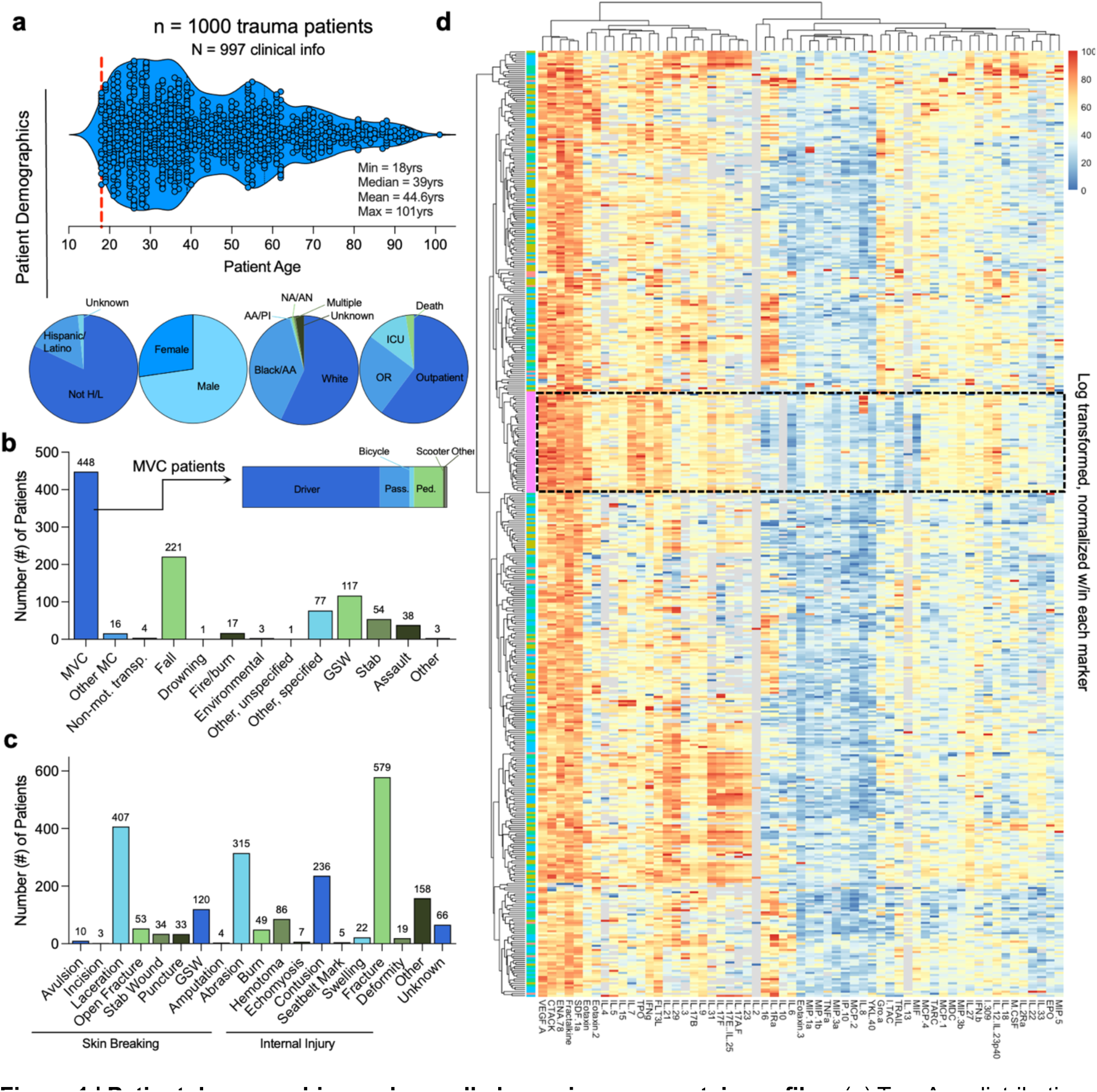
| Patient demographics and overall plasma immune protein profiles. (**a**) Top: Age distribution of trauma patients. Min = 18 yrs, Max = 101 yrs, Median = 36. Bottom: Demographics of patients and trauma outcome. Biologic sex, patient-reported race & ethnicity, trauma outcome. (**b**) Source of trauma. (**c**) Type of trauma. (**d**) Heatmap of protein profiling in N = 1000 trauma patients and N = 50 healthy controls (HC). HC are outlined in black dashed box; data are analyte concentration log transformed and normalized (0 to 100) within marker.

### Significant immunosuppression and select protein upregulation in trauma patients

After normalization and clustering of all proteins, the healthy controls clustered together in a hierarchical dendrogram in comparison to trauma samples (**Figure 1d**, **Supplemental Figure 2**). Tobit regression revealed that traumatic injury led to overall increased levels of 9 proteins (IL-10, MIF, IL-29, TRAIL, IL-23, IL-16, IL-6, IL-1Ra, MIP-5) and decreased levels of 23 proteins compared to healthy controls (MCP-4, IP-10, MDC, FLT3L, Eotaxin-3, CTACK, TARC, MCP-1, Eotaxin, IL-3, TPO, MIP-1b, ENA-78, MIP-3b, I-309, IL-21, IL-12/IL-23p40, IL-17E/IL-25, MIP-1a, VEGF-A, IL-7, IL-8, IL-2, **Figure 2a,b**, **Supplemental Figure 3a**). The greatest differences in protein concentration between the total trauma population and healthy controls were a strong upregulation of IL-10 (23.1% increase versus healthy control, 95% CI: 5.0 – 106.8) and a strong downregulation of IL-2 (97.8% decrease, CI: 91.8 – 99.4). The smallest increase detectable was a 1.46% increase in MIP-5/CCL15 (1.1 – 1.9) and the smallest decrease was a 29.1% decrease in MCP-4/CCL13 (14.4 – 41.3).

**Figure 2.**
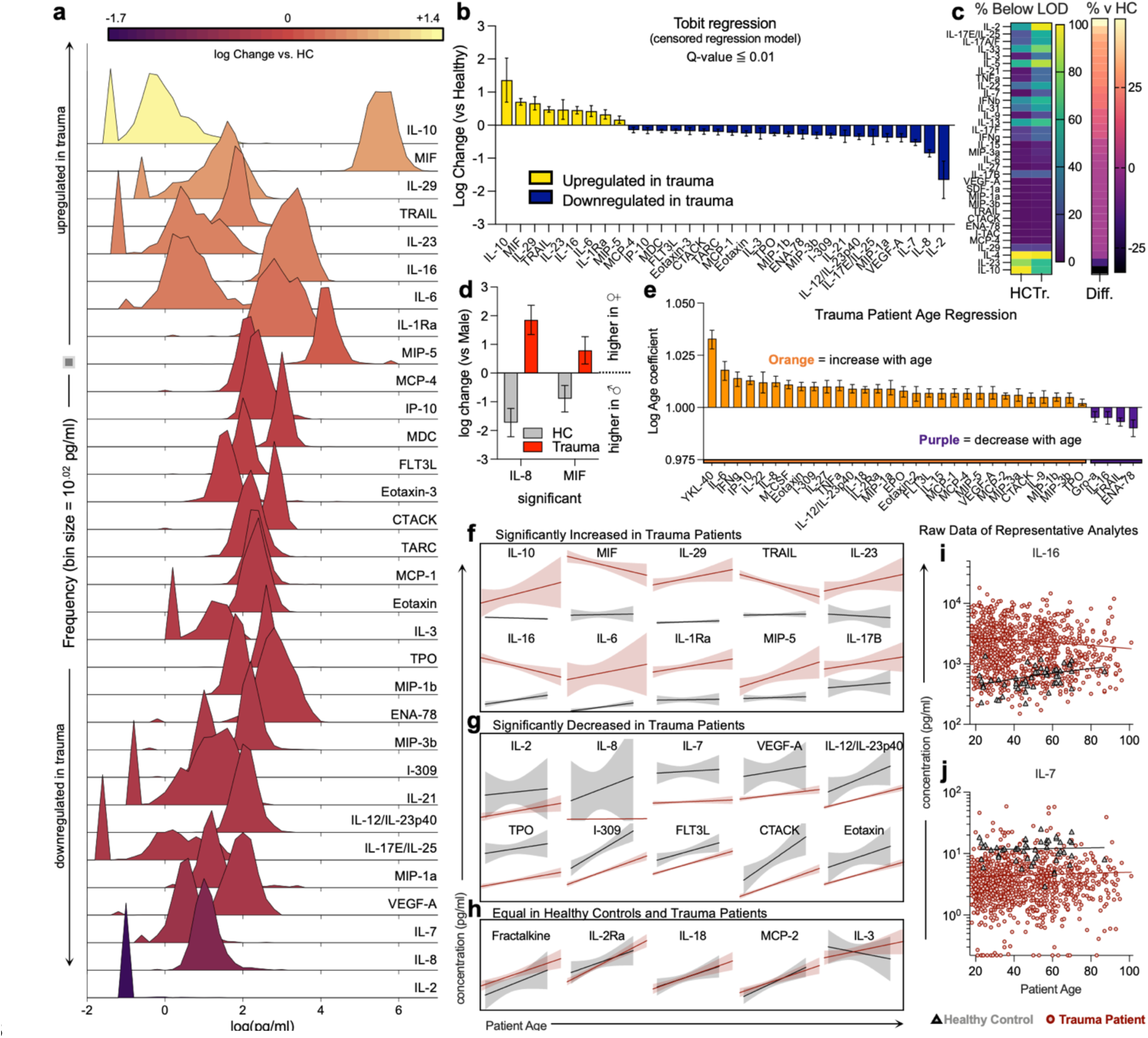
| Differential immune responses in trauma patients are altered by age and sex of patient. **(a)** Distribution of significantly altered proteins in trauma patients compared to healthy controls. (**b**) Tobit regression of significantly altered proteins. Yellow = increased in trauma, Blue = decreased in trauma. Data are log fold change (versus healthy control, HC). (**c**) Proteins that are different in percent (%) below limit of detection (LOD) in trauma patients versus HC (left) and % difference between HC and Trauma (Tr). (**d**) Sex dimorphisms in trauma patients (grey = HC, red = trauma). (**e**) Age coefficients of proteins tested in trauma patients (orange = positive correlation with age, purple = negative correlation with age). (**f-h**) Examples of simple linear regression of age and protein concentration for markers significantly (Tobit) (**f**) increased, (**g**) decreased, (**h**) or equal in trauma patients compared to HC. Raw data of representative analytes that are (**i**) increased and (**j**) decreased in trauma patients compared to healthy control. (**f – j**) Red = trauma patient, black/grey = healthy control. Error bars = with 95% confidence intervals.

Most cytokines and chemokines were detectable (**Figure 1d**, **Supplemental Figure 4**), although several, including IL-10, IL-2, IL-5, IL-23, and IFNβ, were below the assay limit of detection (LOD) in >75% of either trauma or control samples. We therefore evaluated whether trauma influenced the probability of a protein concentration being below the LOD. We found that 30 proteins were significantly more likely to be below LOD in trauma samples versus controls (CTACK, ENA-78, IFNβ, IFNγ, I-TAC, IL-2, IL-3, IL-5, IL-7, IL-9, IL-13, IL-15, IL-17A/F, IL-17E/IL-25, IL-17F, IL-21, IL-22, IL-31, IL-33, MCP-4, MIP-3a, IL-6, IL-27, IL-17B, MIP-1a, MIP-3b, SDF-1a, TNFa, TRAIL, VEGF-A) while 4 were significantly more likely to be below LOD in healthy controls (IL-4, IL-10, IL-23, IL-29), with the largest differences of each being IL-2 (46.3% difference below LOD in trauma vs healthy, p<0.001) and IL-10 (32.7% difference in trauma vs healthy, p<0.001) (**Figure 2c**, **Supplemental Figure 4**). In terms of patient sex, the pattern of differences in protein concentration were similar when controlling for sex in trauma patients versus healthy controls (**Supplemental Figure 5**); however, two proteins had significantly different patterns in males and females. IL-8 and MIF were higher in female trauma patients when compared to male but the opposite in healthy controls **(Figure 2d).**

As patient age spanned 83 years, we evaluated protein levels as a function of age. When controlling for age, trauma patients and healthy controls had a similar pattern of protein concentrations (**Supplemental Figure 5**). Looking at only the population of trauma patients, the concentrations of several proteins, including YKL-40 (CHI3L1), IL-6, IL-8, IFNγ, and others, were positively correlated with patient age **(Figure 2e).** Comparatively, few protein concentrations decreased with age (ENA-78 (CXCL5), TRAIL, IL-16, and Gro-a (CXCL1)). While heterogeneity in protein concentration and a smaller sample of healthy controls prohibited a direct statistical comparison of the regression coefficients between healthy controls and trauma patients, we did observe some age-related patterns in proteins that were significantly different in trauma patients versus healthy controls overall (**Figure 2f – j**). In proteins that were significantly higher in trauma patients versus controls, increasing age had a greater impact on protein concentration (in either direction) (**Figure 2f**). In proteins significantly decreased in trauma patients, the association with age appeared to be more pronounced in healthy controls (**Figure 2g**). The effect of age appeared negligible when protein concentration was equal in healthy controls and trauma patients, with both groups having similar age trends (**Figure 2h**). Raw data for representative analytes is shown in **Figure 2i** and **2j**. Of the proteins that exhibited significant changes as a result of traumatic injury, some variability was observed with self-reported race (IL-12/IL-23p40 and ENA-78, **Supplemental Figure 6**).

### Trauma level, location, and tissue injured alter immune profile

Each patient was categorized as level of trauma activation 1 – 4 per the admitting trauma center, with 1 being the most severe (n = 177) and 4 being alert (n = 490) with a higher risk of death in Level 1 patients compared to level 4 (**Supplementary Figure 7a-g**). Several proteins were significantly associated with trauma level. Specifically, IL-1Ra and IL-16 were increased in severe trauma compared to alert patients (**Figure 3a**) and TPO, IL-21, IL-22, IL-17F, IL-31, IL-33, IL-17A/F, IL-17E/IL-25, and IL-23 were decreased in severe trauma (**Figure 3a**). While not correlated with trauma level, an increase in IL-29 in patients who survived trauma was noted and when compared to IL-10 as a ratio of concentrations, high risk patients could be identified with a low IL-29:IL-10 ratio (**Figure 3b**, **Supplementary Figure 7h-k**). Analyzing these data using a receiver operating curve (ROC) we found an area of 0.933 with a sensitivity of 100% (95% CI: 70.09 - 100%) and specificity of 85% (81.3 – 88.76%) suggest a potential diagnostic tool that can be used to identify patients that have sustained less severe trauma but are at risk for death (**Figure 3c**).

**Figure 3.**
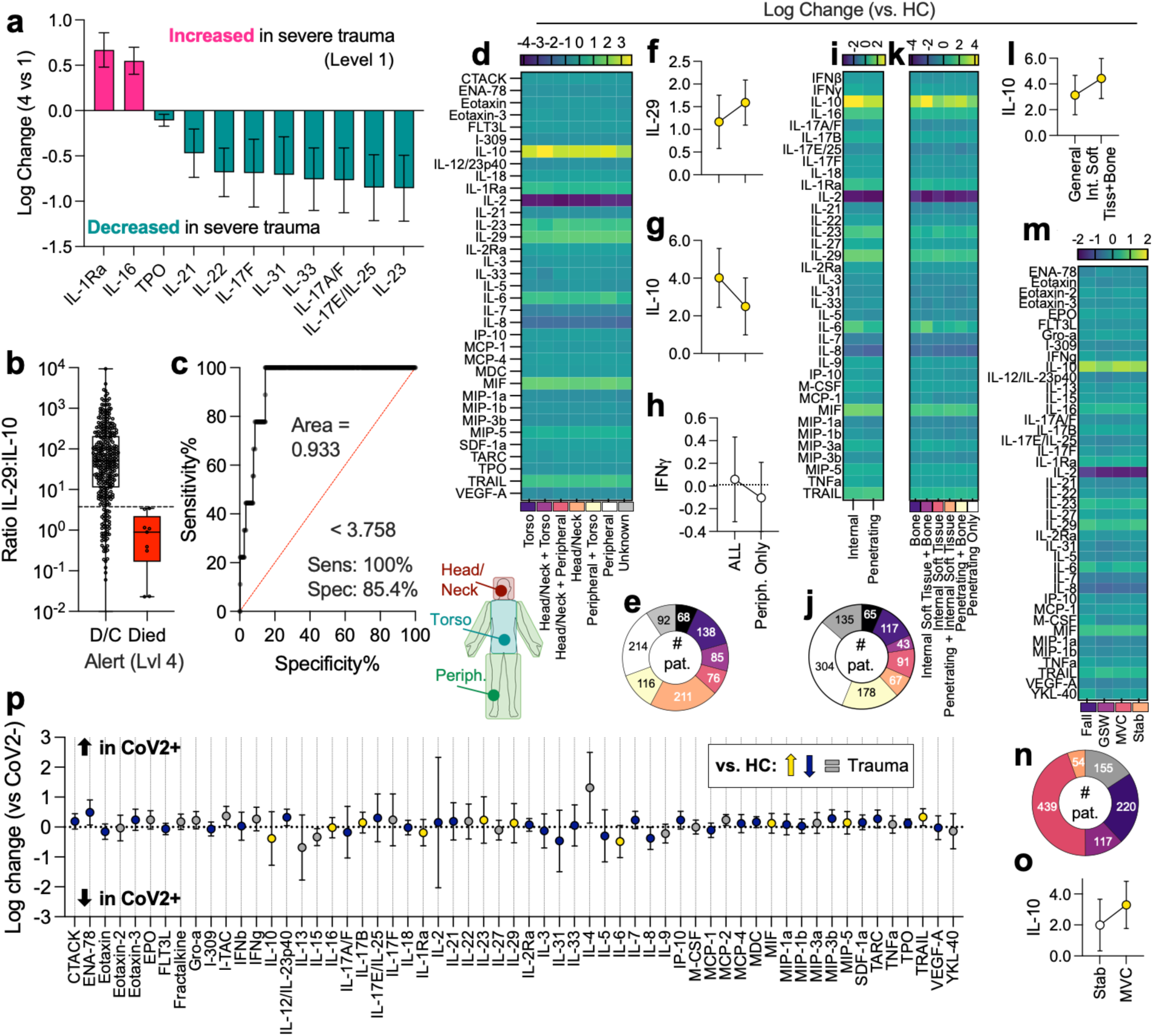
| Trauma level, location, and co-incidence of viral infections cause alterations in immune profiles and reveal IL-29 as a a new predictor of patient survival. (**a**) Protein differences in severe (1) vs less severe (4) traumas. (**b**) IL-29 to IL-10 ratio in survivors versus those that died from less severe trauma. (**c**) ROC of IL-29:IL10 ratio as a predictor of patient survival. (**d**) Log change of protein versus healthy control (HC) by injury location. Purple = decreased in trauma, Yellow = increased. (**e**) Incidence of injury locations. (**f-h**) Cytokine concentration in patients with injuries in all locations versus peripheral only for (**f**) IL-29, (**g**) IL-10, and (**h**) IFNγ. Data are log change ± 95% CI. (**i**) Patients with internal vs penetrating injuries. (**j**) Incidence of injury types. (**k**) Log change by injury types. (**l**) IL-10 change compared to HC in all injuries vs patients with internal soft tissue injury + bone injury. (**m**) Protein profile by trauma source. (**n**) Incidence of trauma source. (**o**) IL-10 change vs HC in stab vs MVC patients. (**p**) Log change in SARS-CoV-2+ trauma patients vs SARS-CoV-2-patients. Yellow = increased in general trauma patients vs HC, Blue = decreased in general trauma patients compared to HC, grey = proteins not significantly different between trauma patients and HC. Data are log change ± 95% CI.

In addition to these differences in trauma level, we saw alterations in immune profile dependent upon the injury location (**Figure 3d,e**). Trauma location was classified as torso only, head/neck only, peripheral only, a combination, all, or unknown (**Figure 3e**). When evaluating IL-29 (an IFNλ) as a function of injury location, we found that patients that had sustained injuries in all locations (torso, head/neck, peripheral) had lower concentration when compared to those that only sustained injuries on the periphery (**Figure 3f**). As with patient survival, this pattern was inverted with IL-10 (**Figure 3g**). There was no effect with IFNγ (which was also not significantly increased in trauma patients) suggesting this is not a pan-interferon pattern (**Figure 3h**).

Variations could also be detected by mechanism of injury, whether or not it was an internal versus penetrating wound (**Figure 3i**), as well as subset by hard (bone) and soft tissue injuries (**Figure 3j,k**). These subsets were selected to enable analysis of variables that are known to affect preclinical models. As with other variables, IL-10 concentration depended upon tissue type, with combination internal soft tissue and bone injuries trending higher than bone wounds alone (4.4% higher than HC versus 2.5%, **Figure 3l**). This correlated with injury mechanisms that were more likely to induce compound injuries such as MVCs (**Figure 3m,n**) wherein IL-10 trended higher than injuries with just one factor (e.g. stab, 3.3% versus 2.0%, **Figure 3o**).

### Immune response to trauma is altered in the presence of active respiratory virus infection

Given sampling occurred during the COVID-19 pandemic, a proportion of patients were tested for COVID-19 upon admission to the hospital (66%). No control samples were tested for SARS-CoV-2 and were excluded from this analysis. Using the data from patients with test results, we compared the immune profiles of trauma patients that tested positive and those that tested negative for active SARS-CoV-2 infection (**Figure 3p**). A total of 25 people tested positive for SARS-CoV-2 and were compared to 612 people who tested negative for SARS-CoV-2 infection. Those without COVID testing or with undetermined results were excluded from this analysis. SARS-Cov-2 positivity appears to be associated with a slight decrease in IL-10 and IL-6 (**Figure 3p)** which were both upregulated in trauma patients relative to HC overall **(Figure 2b).** IL-4, a protein associated with type-2 immunity, trends higher in SARS-CoV-2+ patients, while IL-13 trends lower in SARS-CoV-2+ patients. However, these changes were not statistically significant after correction for multiple comparisons.

### Generation of a predictive equation for patient survival using a subset of four analytes

Given our observation that a novely described responder to trauma (IL-29) could be used in a predictive manner for patient survival in non-severe patients, we investigated further the potential to generate predictive calculations for both non-severe and other trauma patients. When comparing concentrations of proteins in trauma patients that survived to discharge (D/C) versus those that succumbed to their injuries (deceased), a number of proteins trended higher or lower in concentration in the latter group **(Figure 4a)**. Several stood out with stark differences including IL-6 **(Figure 4b)** and M-CSF **(Figure 4c)** that were higher in those that died, whereas IL-29 **(Figure 4d)** and IL-21 **(Figure 4e)** were higher in those that survived. As we had found that a ratio of IL-29 to IL-10 had been predictive of survival, we compared ratios of all cytokines to each other and found that several of the protiens that were highlighted in overall differences between D/C and deceased patients appeared in these estimates (ex. IL-6, **Figure 4f**). Using Random Forest machine learning several of these appeared as having high importance in distinguishing a D/C blood profile from deceased (**Figure 4g**). Isolating proteins that appeared in both machine learning and ratio-based analyses, we generated ROCs of proteins that appeared predictive of survival (IL-6, VEGF-A, IL-10, IL-29, M-CSF, and IL-21) and generated ROC areas that ranged from 0.5049 (non-predictive) to 0.8677 (highly predictive, **Figure 4h**). The variability and lower area under the curve (AUC) values of M-CSF led us to create an abridged list of 5 proteins (IL-6, VEGF-A, IL-10, IL-29, and IL-21) for further analysis. Different machine learning methods generated ROC areas ranging from 0.7600 (XGBoost with class weights) to 0.8957 (Gradient Boost with class weights) with algebraic approaches including regression models ranging from 0.8638 – 0.8873 (**Figure 4i**, **Supplemental Table 5**). To optimize predictive power, we generated an algebraic equation that involved these five top proteins that we called “Vital Injury Protein Evaluation for Recovery” (VIPER, **Figure 4i,j**). VIPER scores were generated for all patients and subset into both Level 1 (most severe) and Level 4 (least severe) groups then evaluated by ROC (**Figure 4k**). While most predictive for survival in least severe patients (0.9695, 0.9310 – 1.000) which is beneficial due to the low alert level for trauma centers of potential life-threatening injuries, the model remained predictive for all patients, with Level 1 having the most variability but still having an ROC of 0.8431 (0.7375 – 0.9486) (**Figure 4l**).

**Figure 4.**
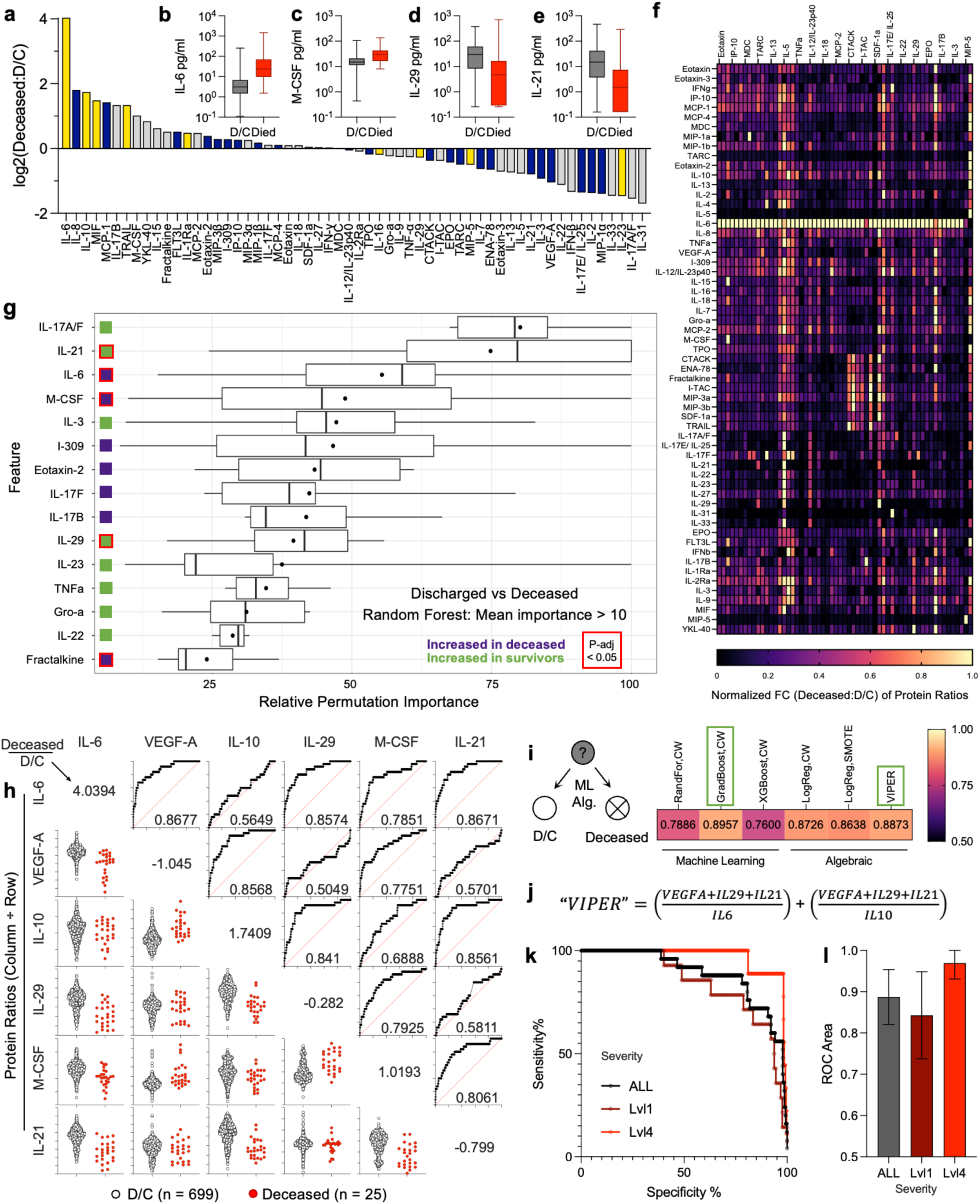
| Development of VIPER equation for survival prediction. (**a**) Fold change of concentration in those that died over those that were discharged home (D/C). (**b – e**) Cytokines altered in moribund patients. (**f**) Fold changes in the calculated ratios of each protein to every other protein for moribund versus D/C. (**g**) Random Forest machine learning permutation importance. Green squares = increased in D/C, Purple squares = increased in deceased, red outline = p < 0.05 when comparing concentrations via Mann Whitney with post-hoc correction for multiple comparisons. Data are mean (dot) and median (line) ± IQR. (**h**) Ratios and ROC curves among six selected important proteins. (**i**) AUC values for multiple machine learning and algebraic models applied to predict patient survival. (**j**) “Vital Injury Protein Evaluation for Recovery” (VIPER) algebraic equation developed using 5 proteins with predictive value for patient death. (**k**) ROC curve of VIPER equation for all patients, and those with level 1 or 4 injuries. (**l**) AUC for VIPER.

### Animal model correlates with human immunomodulation after traumatic injury

As traumatic injuries cannot be randomized in human clinical studies, many researchers rely on animal models to evaluate therapeutics for wound management and tissue reconstruction. To assess similarities and differences between human trauma response and animal models we evaluated RNAseq data following traumatic soft tissue injuries in three common model organisms: mice, rats, and pigs. When mining bulk RNAseq data,^10–13^ we found that early responses to volumetric muscle injury in rats led to increases in *Il10*, *Il16*, *Il6*, *Il23a*, and *Tnfsf10* (encoding TRAIL) similar to human responses (**Figure 5a**).^10,14^ We also found a modest increase in *Mif* and *Il17b*. These trends were maintained in a later timepoint of a porcine volumetric muscle loss (VML) model,^11,15^ however further study of earlier timepoints with increased replicates is needed (**Figure 5b**). This was recapitulated to some degree in freeze-based muscle injuries in mice,^12,16^ including upregulation of *Il10*, *Il1rn* (not seen in rat VML), *Il16*, *Il6*, but with minimal upregulation *Il23a* and *Tnfsf10* (**Figure 5c**). In skin injuries of mice,^13,17^ a robust upregulation of *Il23a* was seen in multiple injury locations including the abdomen, back/dorsum, and cheek at 3 days post-injury (**Figure 5d**).

**Figure 5.**
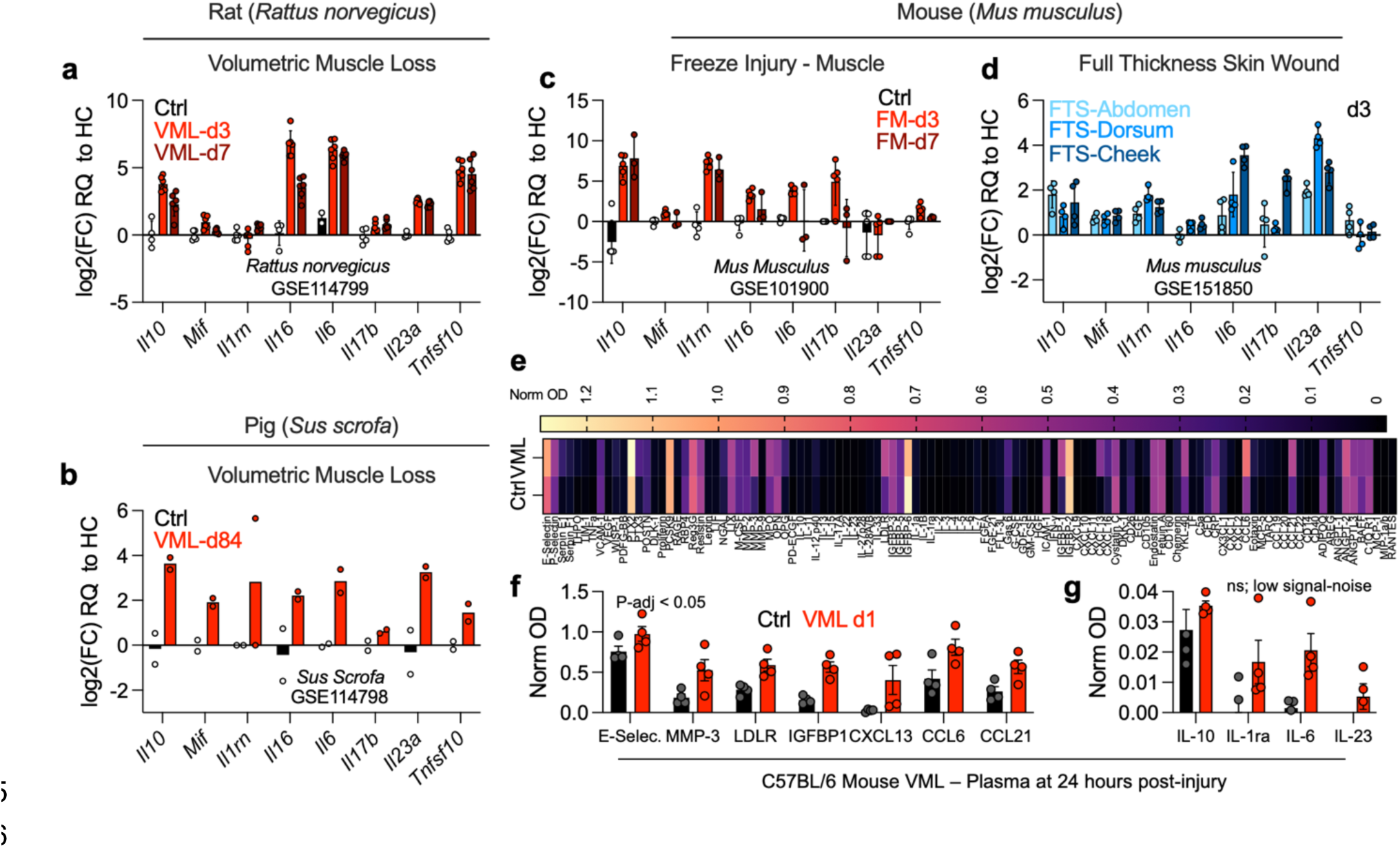
| Comparative animal models for trauma study with human injury response evaluation. (**a-d)** RNAseq data mining for human trauma upregulated markers in local tissue from (**a**) Rat volumetric muscle loss (VML), n = 4 - 6; GSE114799,^14^ (**b**) Pig volumetric muscle loss (VML) n = 2; GSE114798,^15^ (**c**) Mouse freeze injury of muscle (FM), n = 3 - 5; GSE101900,^16^ and (**d**) Mouse full thickness skin wound (FTS), n = 4; GSE151850.^17^ Data are fold change over uninjured/sham controls ± SD. (**e**) Mean normalized optical density (OD) from mouse cytokine/chemokine profiling blot of plasma at 24 hours post VML, n = 4. (f) Values from (**e**) that are significantly different from control (p-adj < 0.05, ANOVA with Tukey post-hoc correction for multiple comparisons). (**g**) Values from (**e**) that were present in human dataset but had low signal to noise ratio (low concentration) in mouse blot. Data are mean ± SEM, n = 4.

Additionally, we performed VML surgeries on mice and evaluated peripheral blood at 24 hours post-injury for immediate responses to trauma that are detectable in the plasma of mice (**Figure 5e-g**). Using a cytokine/chemokine blot to screen these responses, we found multiple proteins that were modulated after injury (**Figure 5e**). Several were systemically altered including chemokines CXCL13, CCL6, and CCL21 (**Figure 5f**). Compared to our human data, we found a low signal to noise ratio (suggesting low concentrations), but trending increases in IL-10, IL-1ra (encoded by *Il1rn*), IL-6, and IL23 (**Figure 5g**). Unfortunately, IL-29 is a pseudogene in mice and rats and could not be evaluated mechanistically, highlighting a limitation in animal models for studying trauma immunology.

### Protein profile classification using machine learning

Through t-stochastic neighbor embedding (tSNE), an unsupervised clustering method, using only the 59 protein concentrations, we identified an island of healthy controls that clustered separate from trauma patients (**Figure 6a**). While some regions had increased enrichment for various factors such as sex, age, outcome, and injury mechanism, only healthy controls could be easily identified (**Figure 6b**). Interestingly, a small cluster appeared that had significantly higher levels of YKL-40 than the rest of the trauma samples (mean log concentration 17.7 pg/mL vs 12.9 pg/mL, p<0.001) (**Figure 6a-b**, **lower left**), despite YKL-40 not being significantly associated with any trauma characteristics aside from a fall injury (**Supplemental Figure 6**). This small cluster was also older than the total trauma population (median 59 years vs 39 years) which we showed was associated with increased YKL-40 in this population (**Figure 2e**) and has been reported in the literature.^18^

**Figure 6.**
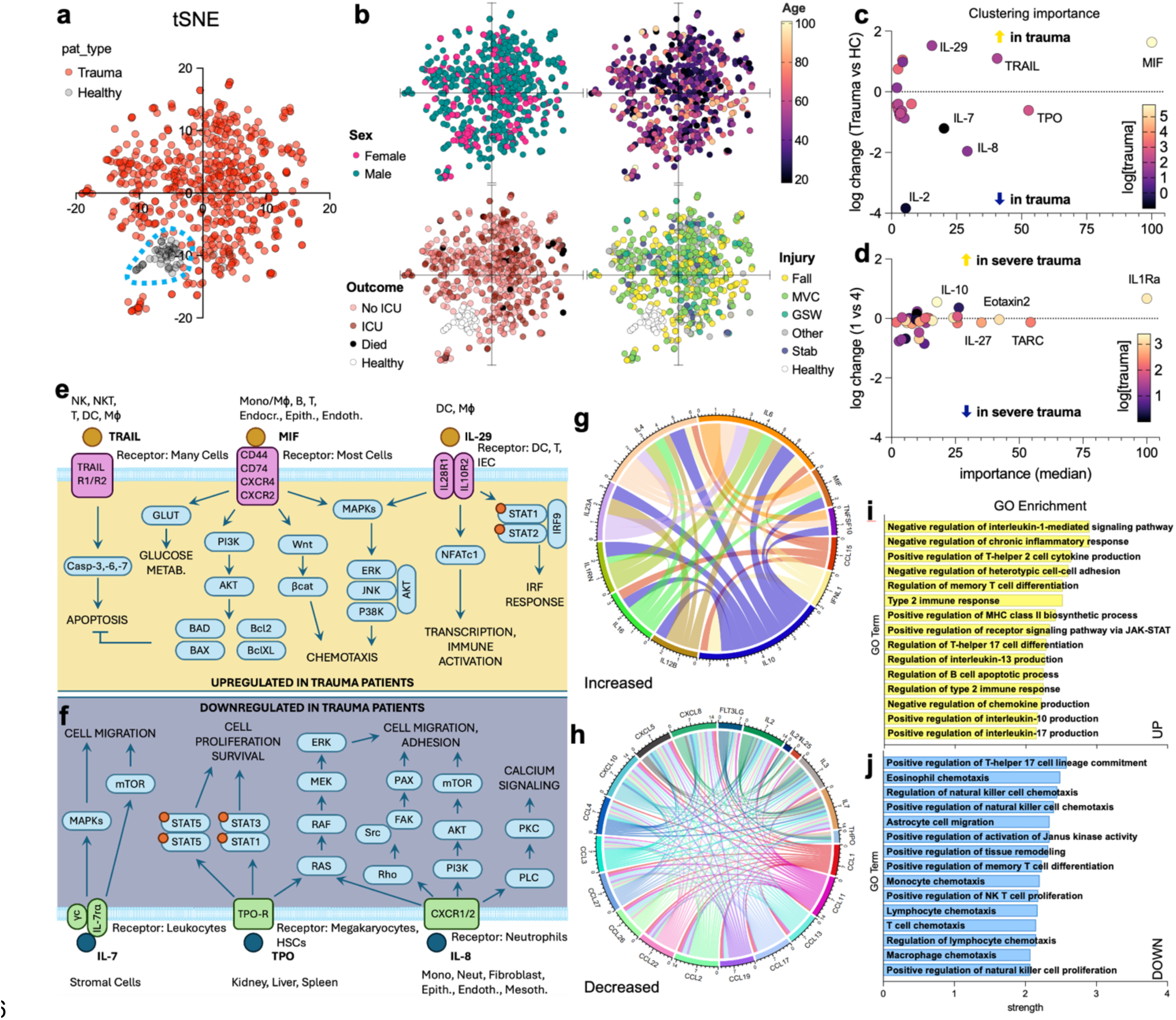
| Machine learning identifies patterns of conserved responses to traumatic injury. (**a**) t-Stochastic neighbor embedding of protein concentrations in trauma patients (red) versus healthy controls (black/grey). (**b**) Labeling of t-SNE diagrams by variables including sex (pink = female, teal = male), age (graded color scale, black = 18yrs, light yellow = 101 yrs), treatment outcome (white = healthy, light pink = outpatient/no ICU, red = intensive care unit (ICU), black = death), and mechanism of injury (yellow = fall, green = motor vehicle crash (MVC), blue green = gunshot wound (GSW), grey = other, blue = stab, white = healthy). (**c-d**) Median importance from machine learning clustering to distinguish (**c**) healthy control from trauma patients, and (d) severe (designation 1) versus alert (designation 4) trauma patients by blood profile. (**e-f**) Proteins (**e**) upregulated or (**f**) downregulated in trauma patients by machine learning and their signaling cascades from receptors. (**g-h**) STRING diagram of interactions of proteins (**g**) increased or (**h**) decreased in trauma patients (determined via Tobit regression). (**i-j**) STRING database gene ontology enrichment of pathways from (**i**) upregulated and (**j**) downregulated proteins

Using random forest, six analytes with the highest relative importance (median permutation importance > 10) for classifying samples as trauma vs control were MIF, TRAIL, and IL-29 (higher in trauma) as well as TPO, IL-8, and IL-7 (lower in trauma) (**Figure 6c**, **Supplemental Figure 9, 7a**). When predicting trauma designations, IL-1Ra, TARC, and Eotaxin-2 were those with the greatest importance in distinguishing lesser (level 4) from life-threatening (level 1) trauma (**Figure 6d**, **Supplemental Figure 10b**).

### Pathway enrichment of systemically altered proteins in trauma patients

Changes to the systemically circulating proteins in trauma patients leads to potential alterations in several signaling cascades and downstream functions (**Figure 6e**). By importing increased (**Figure 6g**) and decreased (**Figure 6h**) proteins into the STRING database, we observed several interactions that could generate a network of different downstream effects. Using gene ontology (GO) enrichment, we found upregulation of pathways associated with the negative regulation of IL-1-mediated signaling pathway and negative regulation of chronic inflammatory response which correlates with general inflammatory immunosuppression (**Figure 1i**). Positive regulation of Th2 cell cytokine production has been associated with wound healing and scar tissue deposition and was also significantly enriched via GO. In decreased protein GO enrichment, we found negative effects on the regulation of TH17 cell lineage commitment, eosinophil chemotaxis, and NK cell chemotaxis (**Figure 6j**).

## DISCUSSION

While there has been significant research into cytokines and chemokines peripherally and within cerebral spinal fluid of traumatic brain injury,^19–23^ less research has been focused on the broad array of other traumatic injuries that present to trauma centers. This observational cohort provided an opportunity to study a large and diverse trauma population, allowing us to characterize how the immune system responds to different trauma-related factors. Some of the proteins we detected have been observed individually in different tissue damage models, but few have been evaluated systemically. The findings present implications for systemic trauma conditions such as multi-organ dysfunction syndrome (MODS).

A robust upregulation of IL-10 was observed in trauma patients as previously described.^2,24^ IL-10 is an immunoregulatory protein that dampens immune responses and inhibits over-activation of immune cells and self-reactivity. We found that IL-10 concentration was dependent upon the location of injury, types of tissues injured, and source of injury. Patients presenting with combined head/neck and torso injuries, those with internal soft tissue and bone injuries, and those injured during a MVC exhibited higher IL-10 levels. These polytrauma patients suggest that core trauma (torso/head), even if not a skin-breaking or penetrating wound, are associated with higher IL-10 and immunosuppression.

### Identification of a new biomarker predictor for patient survival

IL-29 has not been previously associated with traumatic injury and we have shown its robust induction after traumatic injury and increase in patients that survived trauma when compared to those that died. Furthermore, when combining IL-29 concentrations with IL-10 data we developed a ratio that has 100% sensitivity and 85% specificity for patient death after trauma that activated a lower emergency department alert (less severe). Utilization of this ratio could inform patient care by providing a two-analyte panel to predict high risk patients. As IL-29 is an IFNλ that shares the IL-10 receptor beta chain (IL-10RB) with IL-10, the potential opposing roles of IL-10 and IL-29 in post-trauma immunosuppression can be explored to uncover mechanisms of immune response to trauma and unveil new therapeutics. Though IL-22 also utilizes the IL-10RB receptor chain and IFNγ is an interferon, neither had the same pattern as IL-29 or IL-10 suggesting a unique mechanism. If IL-29 is not only a correlate but also a causative agent in patient survival, these insights may yield a useful cytokine-based therapeutic for patients. Unfortunately IL-29 is a pseudogene in organisms used for mechanistic immunology studies and the other IFNλ that is present in mice (IL-28) has unique roles from IL-29. Further work utilizing *in vitro* models may provide some insight into mechanisms, highlighting the need for clinical study to identify mediators of human responses that are lost in preclinical animal modeling. IL-29 is a pleiotropic cytokine that is a player in cancer (regression and persistence), autoimmunity (remission and establishment), and infectious diseases such as COVID-19. This is the first identification of IL-29 having a role in traumatic injury, and this analyte is often left out of standard evaluations making data mining in existing datasets difficult to leverage.

When exploring the data further we found a number of cytokines that were altered in survivors versus those that died. Looking at the relative ratios of proteins, large differences in some protein ratios were observed for those that died compared to those that survived. Combining these results with those of Random Forest machine learning resulted in a list of 6 proteins that were important to distinguishing between survival and death. Through ROC analysis, this list was further refined to 5 proteins (IL-6, VEGF-A, IL-10, IL-29, and IL-21). Applying multiple machine learning models and algebraic methods we developed VIPER, an algebraic equation for the prediction of patient death from this 5-analyte panel. VIPER scores are simple to calculate and have an ROC AUC of 0.8957 across all samples, comparable to the AUC of the machine learning gradient boosting model. Its predictive ability was even stronger for patients that might not be under strict observation for mortality due to lower severity of injuries, with those patients resulting in an ROC area of 0.966. This suggests that VIPER could serve in the future as a clinical tool for predicition of trauma patient death, enabling rapid and increased intervention for patients who are at increased risk of death.

### Novel reporting of trauma-downregulated proteins

Several proteins downregulated in our trauma samples have not been previously reported in the literature in the context of human response to trauma, including IL-17E/IL-25, ENA-78, I-309, and IL-12/IL23p40. Proteins from the IL-17 family are typically associated with autoimmune responses and fibrosis.^25–27^ Here, we observed a previously unreported downregulation of IL-17E/IL-25 in traumatic injury compared to healthy controls. The proportion of samples with undetectable IL-17B was higher in trauma patients versus healthy controls. Patients who experienced either bone wounds alone, or both bone wounds and penetrating soft tissue wounds exhibited a significant decrease in IL-17E/IL-25. These findings suggest the involvement of bone injury in downregulation of IL-17E/IL-25 and possible relationship between severe injuries and prevention of autoimmune related cascades in the early stages of post-trauma responses.

ENA-78 (CXCL5), involved in neutrophil activation and chemotaxis, is upregulated in rat models of hepatectomy^28^ and ischemia reperfusion injury.^29^ We observed a significant decrease in ENA-78 concentration in trauma patients compared to healthy controls. However, those experiencing only internal soft tissue wounds had ENA-78 levels consistent with healthy controls. This suggests that internal soft tissue wounds may not induce robust neutrophil recruitment, possibly because these wounds do not necessarily involve exposure to pathogens. While ENA-78 is involved in neutrophil chemotaxis, I-309 (CCL1) mediates monocyte chemotaxis and was decreased in trauma patients across all wound variables. Additionally, decreases in IL-12/IL-23p40 (promotes macrophage and dendritic cell migration^30^) were observed for all injury mechanisms, wound types, and wound locations. The consistent downregulation of these cytokines across multiple variables indicates a reduction of myeloid migration in the early hours after traumatic injury.

### Novel reporting of trauma-upregulated proteins

Of the proteins we found to be upregulated in trauma, the levels of IL-29 and MIP-5 have not been previously characterized for human trauma patients. IL-29 is known to have antiviral and antitumor properties, and its role in the context of infections has been extensively studied. IL-29 levels have been shown to be elevated in patients with periodontitis,^31^ breast cancer patients with periodontitis,^32^ and those infected with HPV,^33^ and to be decreased in patients with Type 2 Diabetes Mellitus (T2D) and patients with diabetic foot ulcers.^34^ IL-29 was upregulated across all variables, suggesting that IL-29 may be upregulated to prevent the co-incidence of viral or other infections after traumatic injuries. MIP-5, a T-cell and monocyte chemokine that has also been found to be elevated in T2D patients, was also upregulated in trauma patients compared to the control. However, the concentration of MIP-5 was dependent on injury mechanism, with a significantly higher concentration in patients experiencing falls or GSW relative to healthy samples, but not those experiencing MVC or stabbings. MIP-5 was also significantly higher in patients who had combined internal and penetrating wounds, but not for those having either in isolation. Thus, while IL-29 appears to be upregulated in trauma samples across multiple variables, MIP-5 levels appear to depend on specific injury mechanisms. These trends do not appear to be a result of trauma level, as neither MIP-5 nor IL-29 levels were significantly altered in the most severe (level 1) compared to least severe (level 4) injuries.

### Sex and age-based alterations of immunity in trauma patients

Due to the large size of our trauma cohort, we were able to evaluate the correlation of sex and age with immune response to trauma. Patient sex had relatively little association with protein levels with two exceptions: IL-8 and MIF. Female trauma patients had higher levels of these two proteins than male trauma patients, while in healthy females compared to healthy males, these protein concentrations were lower. Sex-specific effects on both IL-8 and MIF levels have been previously reported.^35–39^ Our observation that sex-specific IL-8 and MIF levels in trauma patients exhibit an opposite trend from what is observed in healthy controls indicates that a disruption of the immune system due to traumatic injury can also lead to a disruption of sex-specific immune responses. This should be taken into account for the development of therapeutics that incorporate knowledge of sex-specific immune responses.

Despite the large age range of the trauma patients, we observed significant correlations of age with concentration of proteins including ones previously reported in the literature^18,40–49^ along with several that were not significant in published data^40,50^ but reached significance in ours. Age-related increases in CTACK, I-309, IL-22, IL-2ra, IL-8, IL-9, MCP-2, MIP-1a, MIP-3a, MIP-3b, and MIP-5 have not previously been described. The majority of these proteins are involved in immune cell chemotaxis with gene ontology analysis on proteins upregulated with age having 7 of 15 top enriched pathways involved chemotaxis or cell migration (**Supplemental Figure 11**). ENA-78, Gro-a, IL-16, and TRAIL decreased with age which has not been previously reported. TRAIL induces cell death and can be produced by regulatory T cells that are known to decrease in number with age. ENA-78 and Gro-a both have chemotactic activity for neutrophils, while IL-16 stimulates migration of eosinophils, monocytes, and CD4+ lymphocytes. These observations align with previous reports of attenuated neutrophil and lymphocyte chemotaxis with age.^51–53^ These data further highlight immune complexity where a cascade of proteins that may have overlapping functions can be differentially altered in response to factors such as age or traumatic injury. Additional study is warranted to explore the immunological component to frailty commonly observed following injuries sustained by geriatric patients.

In addition to our primary findings, we also explored several differences of interest for continued investigation. The co-incidence of viral infection in trauma patients was associated with variations of their immune profile. Given the strong induction of proteins that are regulatory or can act in regulatory manners (IL-10, IL-29, IL-6, IL-1Ra), the negative correlation of severe trauma with the ability to fight off infections like pneumonia, and post-acute sequelae of both infections and trauma, these data support future studies on the networked role of responses to pathogens and traumatic injury. Prior studies revealed distinct patterns of inflammatory biomarkers that distinguish blunt trauma patients with nosocomial infections from those without infections.^54^ In addition to infectious disease, a small subset of trauma patients in our cohort reported co-incident cancer which was associated with higher IL-10 levels (**Supplemental Figure 12**). Increased IL-10 has been associated with worse outcomes in both tumor clearance and trauma recovery,^55,56^ highlighting the need for a deeper understanding of how traumatic injuries can differentially impact specific patient populations.

Investigation into the human immune response to trauma yields insight for evaluating downline patient outcomes, understanding the basic biologic principles of the human response to traumatic injury, and identifying targets for therapeutics. In this study we were able to confirm several findings in the literature on the effects of traumatic injury on protein levels, examine factors such as trauma location and trauma level that affected these proteins, and identify new mediators of the systemic human inflammatory response to injury.

While our cohort provides a broad look at various trauma factors, it is important to note some limitations with these data. Convenience sampling of participants from trauma sites could introduce unknown bias into our cohort for which we were unable to control, but it is not possible to do a randomized clinical trial of traumatic injury. As we did not have complete injury severity score (ISS) information for all patients, and thus compared trauma levels. While associated with severity, trauma level (as with other metrics) can be subjective. Despite this, there was a strong correlation between trauma level and risk of death (Trauma Level 1 (0.09, 95 % CI: 0.06-0.016) versus Trauma Level 4 (0.02, 95 % CI: 0.01 – 0.05)). In addition to severity, as this study evaluates acute response to trauma, a focused longitudinal study of timepoints hours, days, and weeks after traumatic injury would provide an understanding of temporal immune dynamics. We were also limited in our ability to independently assess lower incidence trauma (e.g. severe burns) and co-morbidities due to smaller numbers. A deeper evaluation of the different immune responses in non-trauma controls with varied comorbidities (e.g., cancer) and more specific trauma cohorts is needed to to deconvolute the intricate interactions of trauma and human diseases. Despite these limitations, we believe this is largely overcome with the large and diverse cohort.

This study has identified novel trauma-associated immune changes in humans that are altered based on age, sex, trauma source, injury location, and trauma level. These differences can inform future mechanistic studies and clinical evaluations based on the function of different proteins that we have detected. While infectious diseases can take days to mount and potentially cause tissue damage, traumatic injuries cause a large disruption in homeostasis in a matter of seconds. As such, our lab and others have been interested in the prevention of damaging autoimmunity after acute tissue damage caused by traumatic injury. In this study, alterations in TRAIL, IL-29, IL-23, IL-17, IL-1Ra, IL-10, IL-6 and others involved in promoting or inhibiting autoimmune like conditions show a strong connection between immunologic self-reactivity and response to traumatic injury that must be investigated further. Identification of IL-29 as a biomarker for survival from traumatic injury can directly affect clinical care and is a topic for future evaluation, albeit with limitations in mechanistic studies due to the absence of this protein in mice. Potential for IL-29 to be adopted as a therapeutic can also be investigated to determine if this is not only a correlate of survival but also a potential route for intravenous administration in at-risk patients.

## Data Availability

Data and abbreviated clinical information will be made available in supplement after peer review. Not all detailed clinical information gathered during the study will be made available to prevent de-identification of samples and participants. Some values may be changed to ensure privacy of data while maintaining ability to complete any necessary meta-analyses.

## ACKNOWLEDGMENTS

We would like to thank the patients whose samples were used in this study-without their contributions the research would not be possible. We would also like to thank Vanathi Sundaresan for assistance in laboratory organization, Sabrina DeStefano for assistance in collecting mouse blood samples, Andrea Lucia Alfonso for assistance in organizing human samples, and Dr. Daniel Chertow for helpful discussions. This work was funded by the intramural research programs of the National Institute of Biomedical Imaging and Bioengineering, and National Institute of Allergy and Infectious Diseases, National Institutes of Health (NIH). <u>Disclaimer</u>: The contents of this publication are the authors’ sole responsibility and do not necessarily reflect the views, opinions, or policies of the NIH, the Uniformed Services University of the Health Sciences, the Department of Health and Human Services (HHS), or the Department of Defense. Mentioning trade names, commercial products, or organizations does not imply endorsement by the U.S. Government. This work was prepared by a military or civilian employee of the US Government as part of the individual’s official duties and therefore is in the public domain and does not possess copyright protection (public domain information may be freely distributed and copied; however, as a courtesy it is requested that the authors be given an appropriate acknowledgement).

## MATERIALS AND METHODS

### Clinical study and sample collection

Clinical study and sample collection proceeded as previously described^7^ (See Report No. DOT HS 813 399). Briefly, samples were collected by convenience sampling in the time period of May 9th 2020 to January 26th 2021 during an ongoing National Highway Traffic Safety Administration (NHTSA) study of drug prevalence among adult (age 18+) trauma victims who had severe enough injuries for transport by EMS and had a trauma team activated/alerted at selected Level-1 trauma centers. These specimens were collected from patients who were already having blood drawn as part of medical treatment at the trauma centers and were made available for research purposes. Serological analyses were conducted on excess plasma samples from the study when possible.

The study was conducted in accordance with Good Clinical Practice, the principles of the Belmont Report, and HHS regulations enumerated under 45 CFR 46. Five of the sites had the Chesapeake/Advarra Institutional Review Board as the central IRB (Advarra Protocol # Pro00022129), and the Jacksonville, FL site had the University of Florida Institutional Review Board as the IRB of record. De-identified samples and other data were included in the study through IRB-approved waivers of consent and authorization. Medical records or other secondary sources such as emergency medical services run reports and crash reports provided the demographic information. De-identified samples were then sent on dry ice overnight to NIH and stored at −80°C until processing. De-identified healthy volunteer blood samples were collected under clinical protocol NCT0000128 and NIH IRB-approved protocol 99-CC-0168 at the National Institute of Allergy and Infectious Diseases, National Institutes of Health, Bethesda MD.

### Patient and Injury Categorization

Based on clinical data, patients were sorted into a variety of subcategories for further analysis. Patients were sorted into one of five injury mechanisms: fall, gushot/shotgun wound (GSW), motor vehicle crash (MVC), stabbing, or other. Among all injuries, the injury location or locations were also identified for each patient, with injuries categorized as occurring in the head/neck area, torso, peripheral regions, a combination of two of these, or in unknown locations. Regardless of injury mechanism or location, patient injuries were also sorted into the categories of internal (no broken skin), or penetrating (breaking the skin). A more granular understanding of the effects of tissue types on immune response was also obtained by sorting injuries based on tissue involvement, where the major tissue types were categorized as bone, internal soft tissue, and penetrating (breaking the skin), or a combination of two of these. Due to a lack of complete injury severity scores for all patients, the injury severity was analyzed using clinically-assessed trauma levels 1-4 as a proxy, where level 1 was the worst trauma level.

### Electrochemiluminescence detection of protein analytes

Samples were processed and tested on a custom 59-plex MesoScale Diagnostics electrochemiluminescence assay as per manufacturer’s instructions. Protein concentrations were calculated off of standard curves.

### Volumetric muscle loss model

Six (6) week-old C57BL/6J female mice (Jackson Labs) were received and equilibrated in the animal facility for 7 days prior to surgery. The day preceding surgery mice were anesthetized under 4% isoflurane in oxygen and maintained under 2% isoflurane prior to removal of hair from the legs using an electric razor followed by depilatory cream. Remaining depilatory cream was removed with a gauze pad with 70% isoflurane and mice were returned to a clean cage until the following day. The day of surgery mice were anesthetized under 4% isoflurane and maintained under 2% isoflurane diluted in oxygen for the duration of the procedure (roughly 6 minutes). Perioperatively, mice received 0.5 mg/ml Buprenorphine ER (ZooPharm) subcutaneously followed by eye lubricant and surgical site was sterilized with three successive rounds of betadine followed by 70% isopropanol. A 1 cm incision was made in the skin overlying the quadriceps muscle (quad) followed by the underlying fascia. A 30 mg portion of muscle was resected from the midbelly of the quad resulting in a 3mm x 3mm injury. Skin was closed using 3 – 4 wound clips (7 mm, Roboz) and the procedure was repeated on the contralateral leg. Mice were kept warm during the procedure through hand warmers separated by sterile drapes. After surgery, animals received 100 ul warm surgical saline and were monitored under a heat lamp until ambulatory and grooming. All animal research was supervised and approved by the NIH Clinical Center ACUC under protocol NIBIB23-01.

### Mouse cytokine and chemokine protein array

Peripheral blood was collected from the mouse peri-euthanasia through a submandibular bleed. Serum (100 µl) was loaded onto a Proteome Profiler Mouse XL Cytokine and Chemokine Array (R&D Systems) as per manufacturer’s instructions. Briefly, assay membranes were incubated with blocking buffer for one hour before addition of samples. Each sample was diluted to a final volume of 1.5 mL prior to addition, and samples were incubated on the membranes overnight at 4 °C on a rocking platform. Membranes were washed with wash buffer on a rocking platform three times (10 minutes each), and then incubated with the diluted detection antibody cocktail for 1 hour at room temperature on a rocking platform. Membranes were then incubated with horse radish peroxidase-conjugated streptavidin for 30 minutes at room temperature on a rocking platform shaker, washed three times, and incubated with the Chemi Reagent Mix for 1 minute. Images were acquired using a BioRAD Gel Imager, and quantification was conducted using ImageJ.

### STRING Analysis

The lists of upregulated and downregulated proteins were searched for in STRING, a database of known and predicted protein-protein interactions.^57^ The combined score for interactions between pairs of proteins was exported and used to generate the chord diagrams in R. Lists of functional enrichments, in the form of Biological Processes (Gene Ontology), were exported and the 15 pathways with the lowest false discovery rates (FDR) were selected for each list of proteins. These were then sorted by strength.

### Data Analysis and Statistics

Each analyte was subject to lower and upper limits of detection (LOD) of the assay, both for healthy control and trauma samples. Values above the LOD were set to the analyte-specific assay upper LOD + 10%. Values below the lower LOD were set to zero for data visualization and were censored for regression analysis. All protein concentrations were log-transformed.

To assess the relationship between protein concentration and independent variables of interest, we chose to use Tobit regression, which estimates linear relationships when the outcome is censored from one direction, in this case, left censoring at the lower limit of detection. Using the AER package in R,^58^ we constructed individual Tobit models for each of the 59 proteins and the following independent variables: patient type (trauma or healthy control), age (mean-centered), sex, COVID-19 infection status, injury mechanism, trauma level, general and specific wound types, and wound location. Age and sex models controlled for patient type; all others were univariable. Exponentiated regression coefficients and 95% confidence intervals are reported as percent change in protein concentration per unit increase in the independent variable. When reporting whether an association between a protein and predictor was statistically significant, we separately computed q-values for the probability that the association was positive or negative^59^ and controlled the FDR at 0.01 for each of the comparisons.

Exploratory cluster analysis of normalized protein concentrations was conducted using Van der Maaten’s Barnes-Hut implementation of t-Distributed Stochastic Neighbor Embedding (t-SNE) in R using the default settings in the Rtsne package.^60^ Random Forest from the tidymodels^61^ R package was employed to determine the relative importance of features for classifying trauma vs control. All continuous variables (age, protein concentrations) were normalized, patient sex was one-hot encoded. For each variable in the model, we calculated permutation importance across 10 model iterations. Relative importance was determined by dividing each variable’s permutation importance score by the largest importance score of all variables for each of the 10 iterations, then multiplying by 100. See the **Supplemental Materials** and Mayer et al for more information.^62^ Tobit regression, cluster analysis, and random forest analyses were conducted using R version 4.2.x.^63^

IL-29:IL-10 concentration ratio was generated as a ratio (pg/ml:pg/ml) of the two cytokines and the resulting ratio was compared in level 4 trauma activation patients via receiver operating curve (ROC) in GraphPad Prism v10.2.2 comparing those that were discharged to home (D/C, n = 343) versus those that died (n = 9). Patients that were discharged to any other location were excluded from analysis (rehab, correctional facility, hospice, left against medical advice). Any values that were stated as below limit of detection were replaced with the concentration of the lowest point on the standard curve or lowest detected sample, whichever was smaller. Exploratory analysis of cancer data was conducted on GraphPad Prism with undetected values treated as previously stated and comparisons made without corrections due to the exploratory nature of the evaluation.

## DATA AND CODE AVAILABILITY

Statistical comparisons with resulting estimates, p-values, and q-values, along with designation of significance will be made available in supplement along with code used for these analyses (via GitHub) after peer review.

---

**Supplemental Figure 1.**
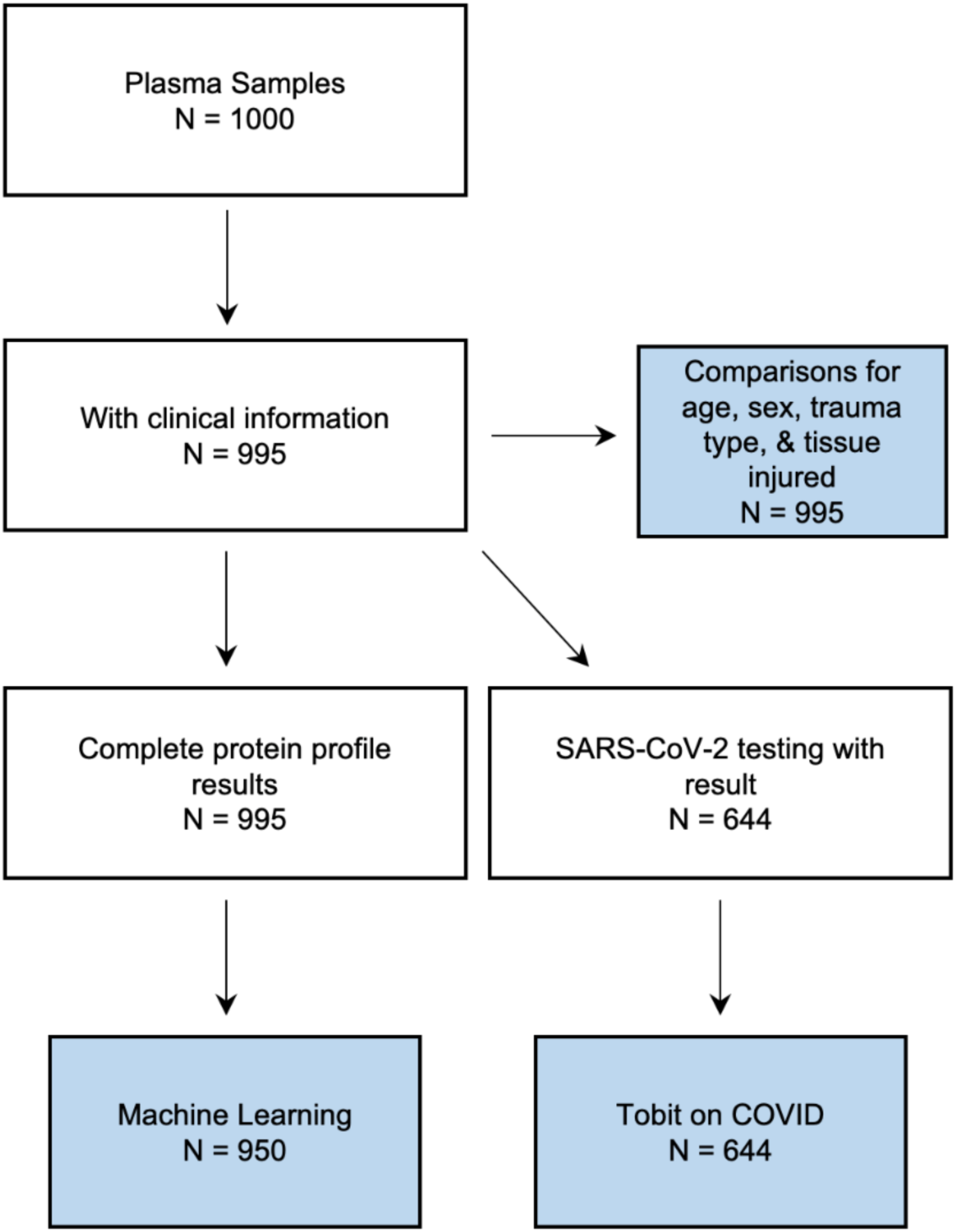
| Sample and data analysis workflow. White boxes = sample input. Blue boxes = analyses.

**Supplemental Figure 2.**
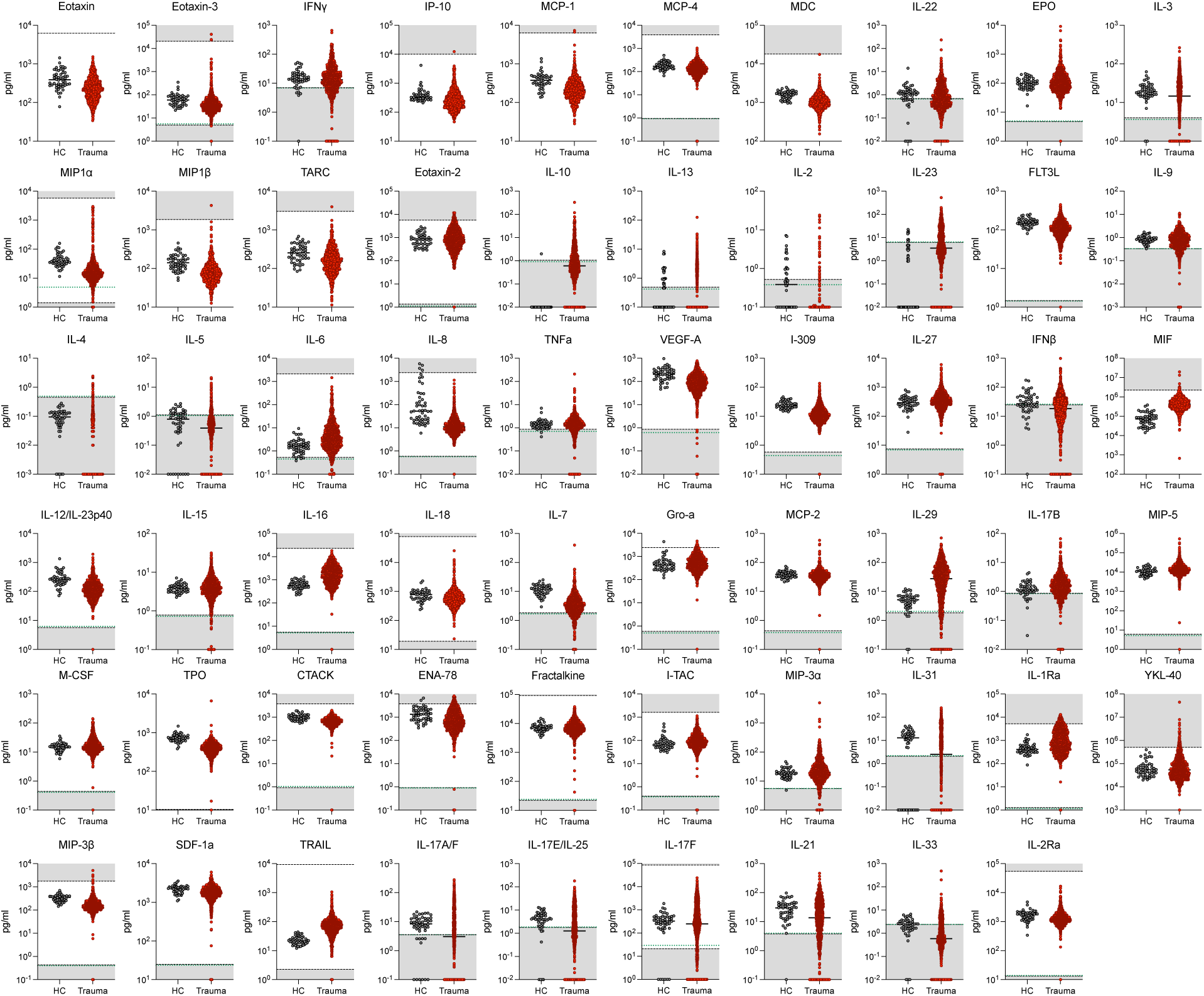
| Raw data display for all analytes measured in full cohort. Grey = Healthy controls (HC); Red Points = Trauma Patients. Green Iline = threshold for healthy control samples, Black line = threshold for trauma samples. Grey shaded areas = above or below highest known standard curve value.

**Supplemental Figure 3.**
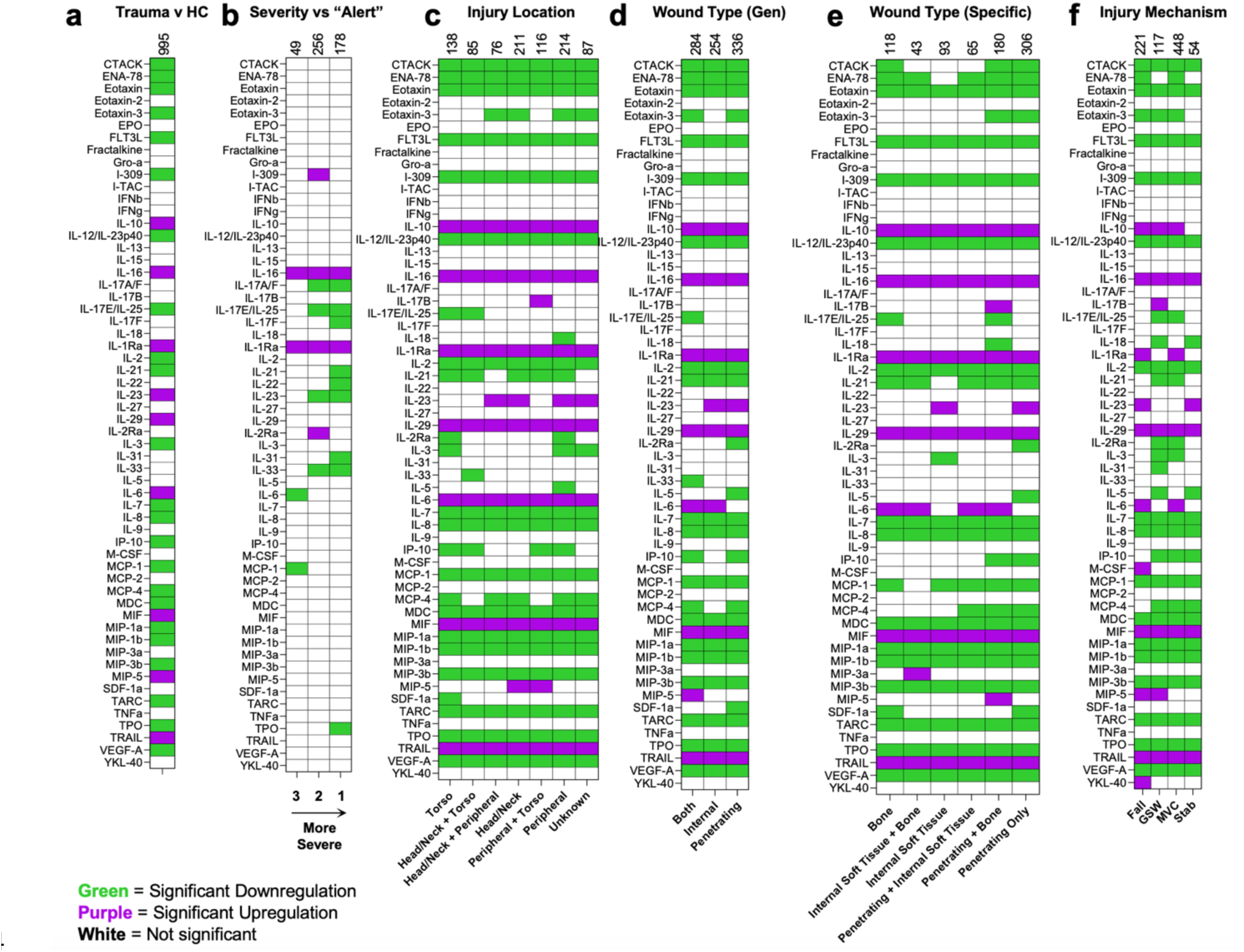
| Binary Heatmaps of Significant Tobit Estimates. Heatmaps of statistically significant Tobit estimates for (a) Trauma versus healthy controls, (b) Different trauma levels compared to trauma level 1, (c) Injury location, (d) general wound type, (e) Specific wound type, and (f) injury mechanism.

**Supplemental Figure 4.**
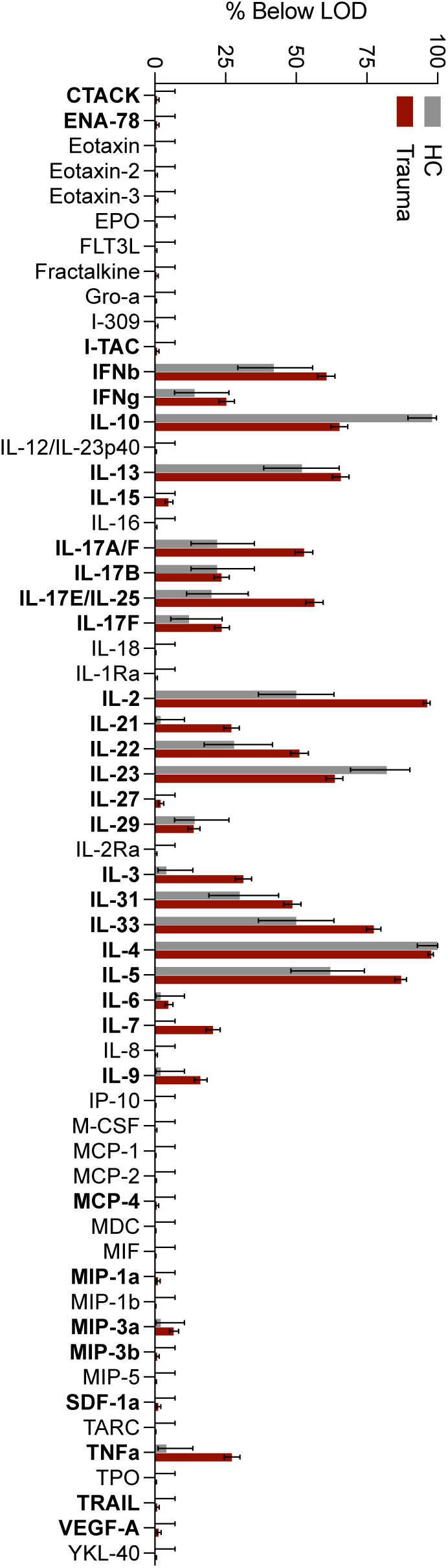
| Proportion below limit of detection. Proportion of samples that were below the limit of detection (LOD) for the assay. Grey = Healthy controls, Red = trauma patients. Data are % below LOD ± 95% confidence intervals (Wilson).

**Supplemental Figure 5.**
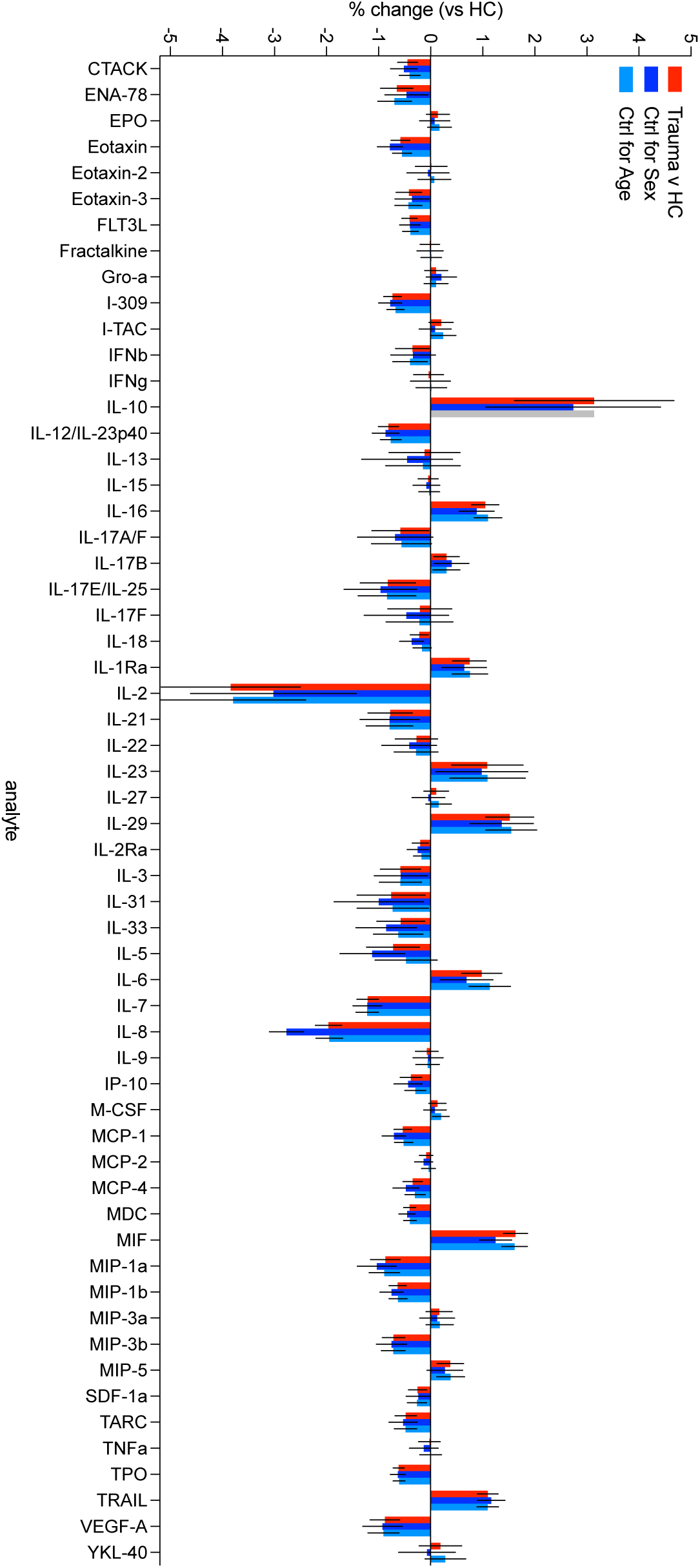
| Estimates of change from healthy volunteers when controlled for sex and age. Tobit estimates for trauma vs healthy controls (HC) with overall (red), controlled for sex (blue), and controlled for age (light blue). Data are log change ± 95% confidence intervals.

**Supplemental Figure 6.**
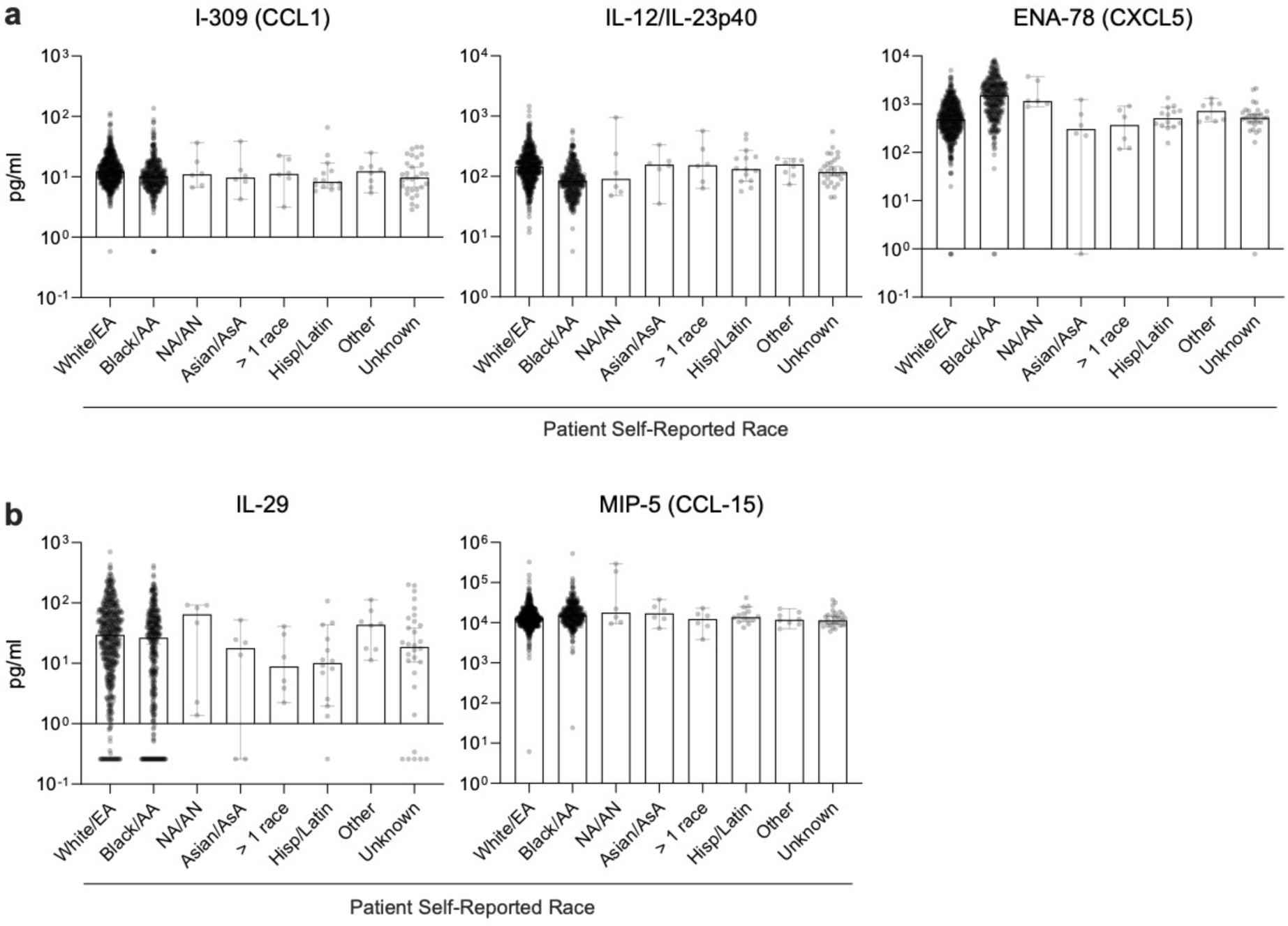
| Effect of self-reported race on concentration of cytokines and chemokines in the blood of trauma patients. (a) Novel downregulated proteins as a function of self-reported race. (b) Novel upregulated proteins as a function of self-reported race. EA = European American, AA = African American, NA/AN = Native American/Alaska Native, AsA = Asian American. Data are mean ± standard deviation.

**Supplemental Figure 7.**
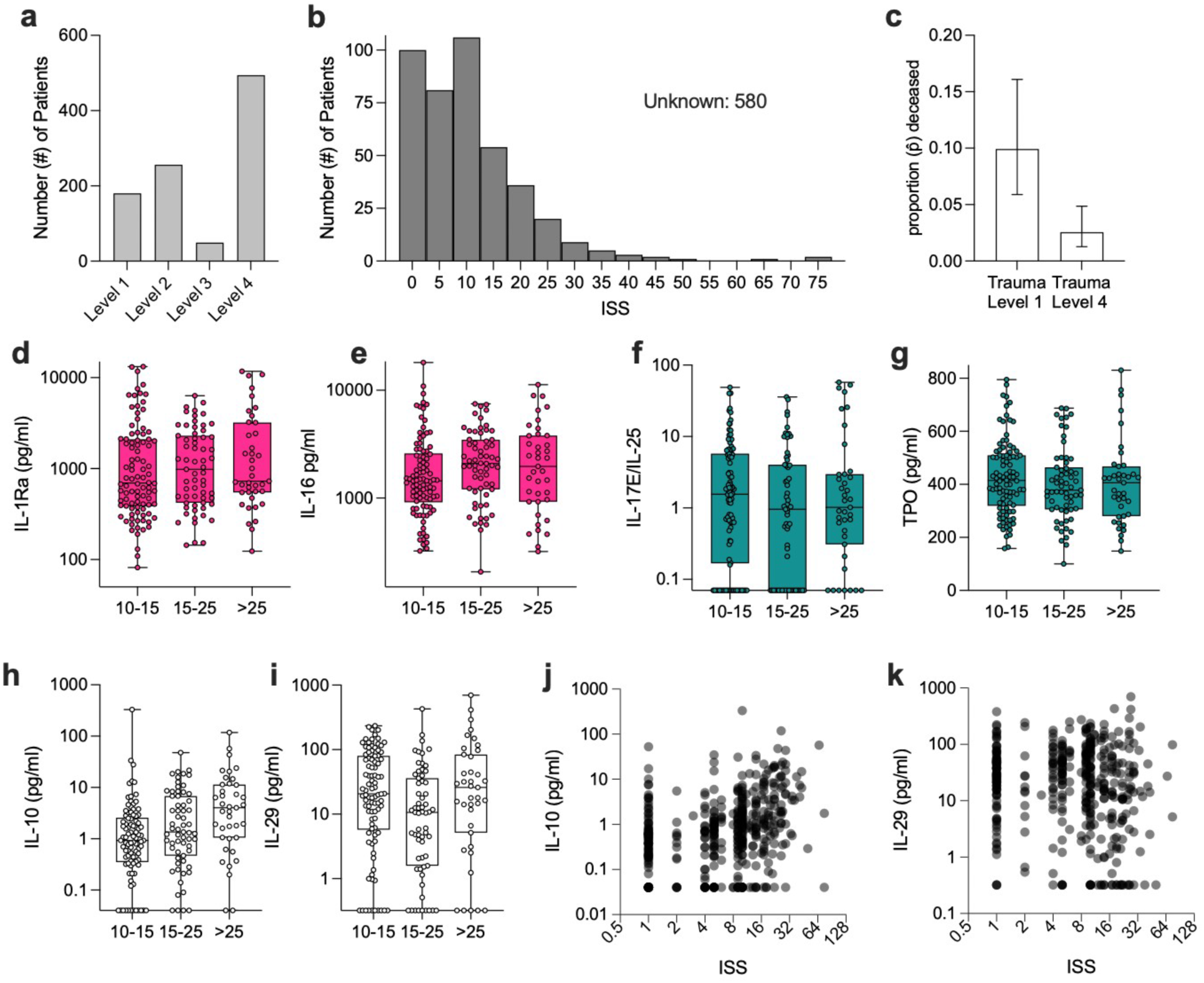
| Trauma level correlates with risk of death and protein concentration patterns are conserved with those that have an injury severity score (ISS). (a) Number of patients categorized into different trauma levels (Level 1 = most severe trauma center activation, Level 4 = least severe trauma center activation). (b) Distribution of ISS including those listed with ISS (including those with one injury). (c) Probability of death when admitted under trauma activation level 1 versus trauma activation level 4. (d – g) Distribution of cytokine/chemokine concentration of upregulated (d, e) IL-1Ra and IL-16 respectively, and downregulated (f, g) IL-17E/IL-25 and TPO respectively, proteins as determined by Level 1 versus Level 4 concentration. (h, i) Concentration of (h) IL-10 and (i) IL-29 by ISS level and (j-k) Correlation of (j) IL-10 and (k) IL-29 concentrations as a function of ISS.

**Supplemental Figure 8.**
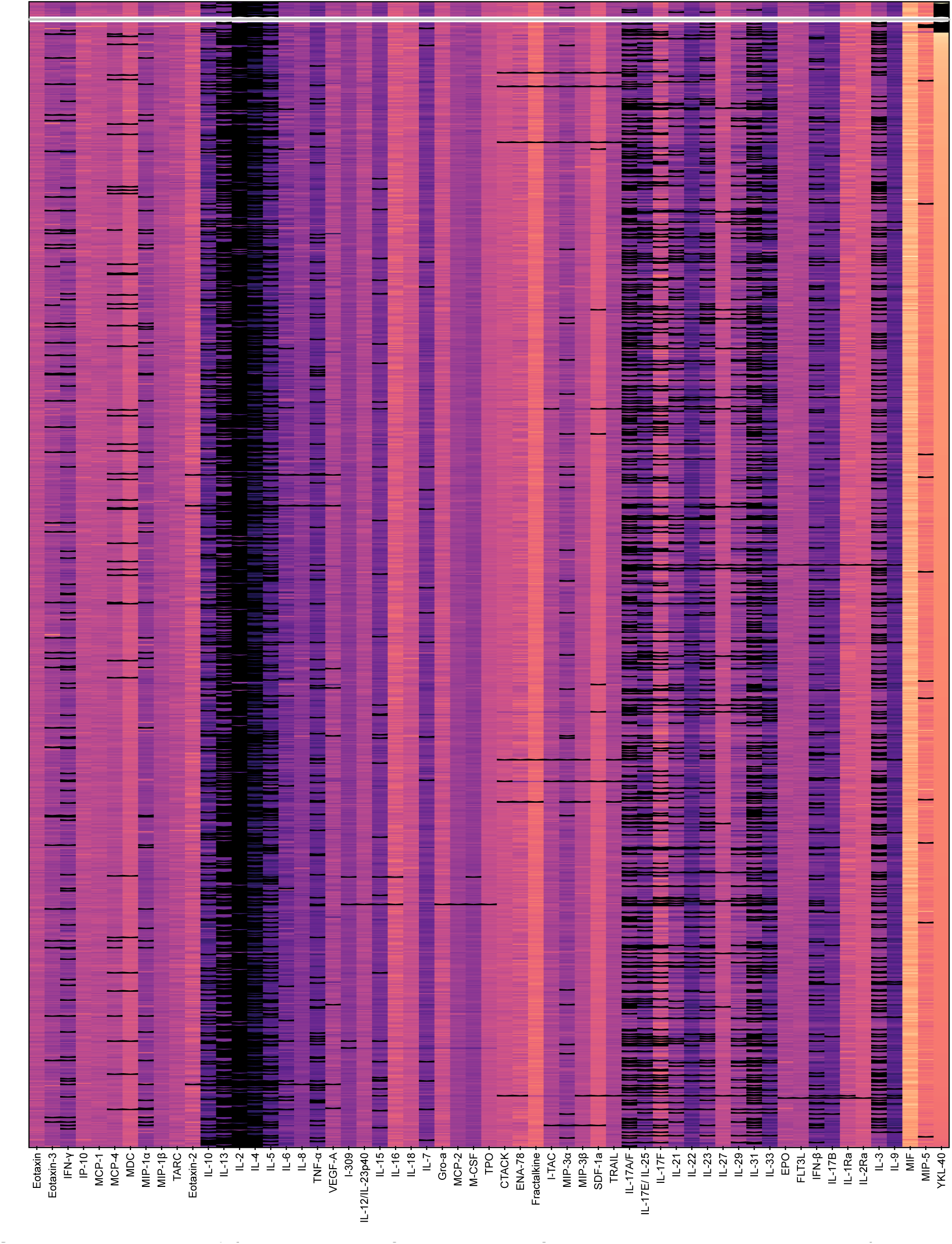
| Phenotypes of sub-cluster of patients with variable cytokine/chemokine levels. Log-normalized concentration (pg/ml), black = above or below limit of detection (LOD).

**Supplemental Figure 9.**
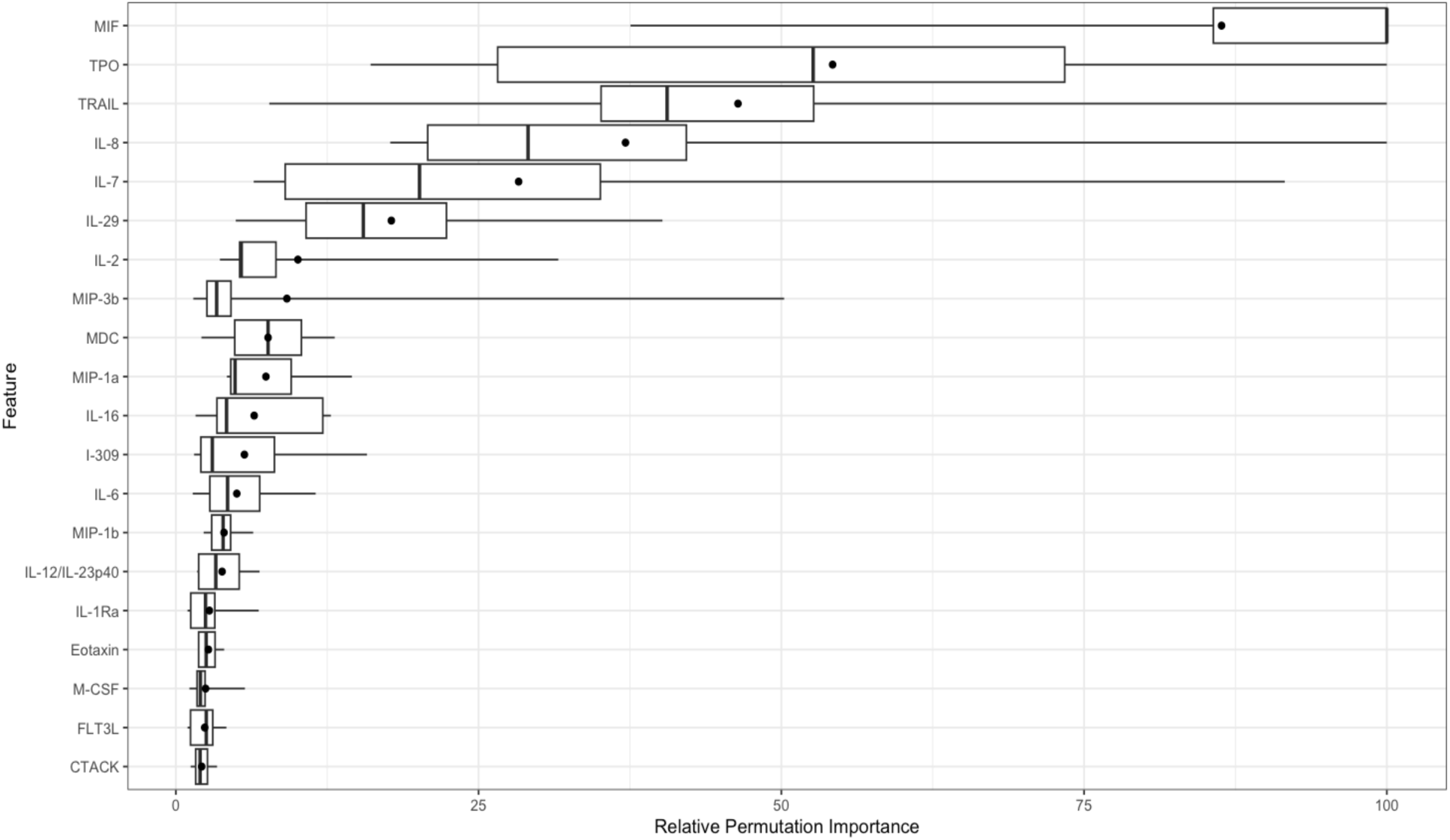
| Relative permutation importance of different proteins on identification of trauma versus healthy control via machine learning. Boxplots are the protein’s minimum, first quartile, median, mean (black dot), third quartile, and maximum relative importance values across 10 model iterations. Importance is ordered by average relative importance across all iterations.

**Supplemental Figure 10.**
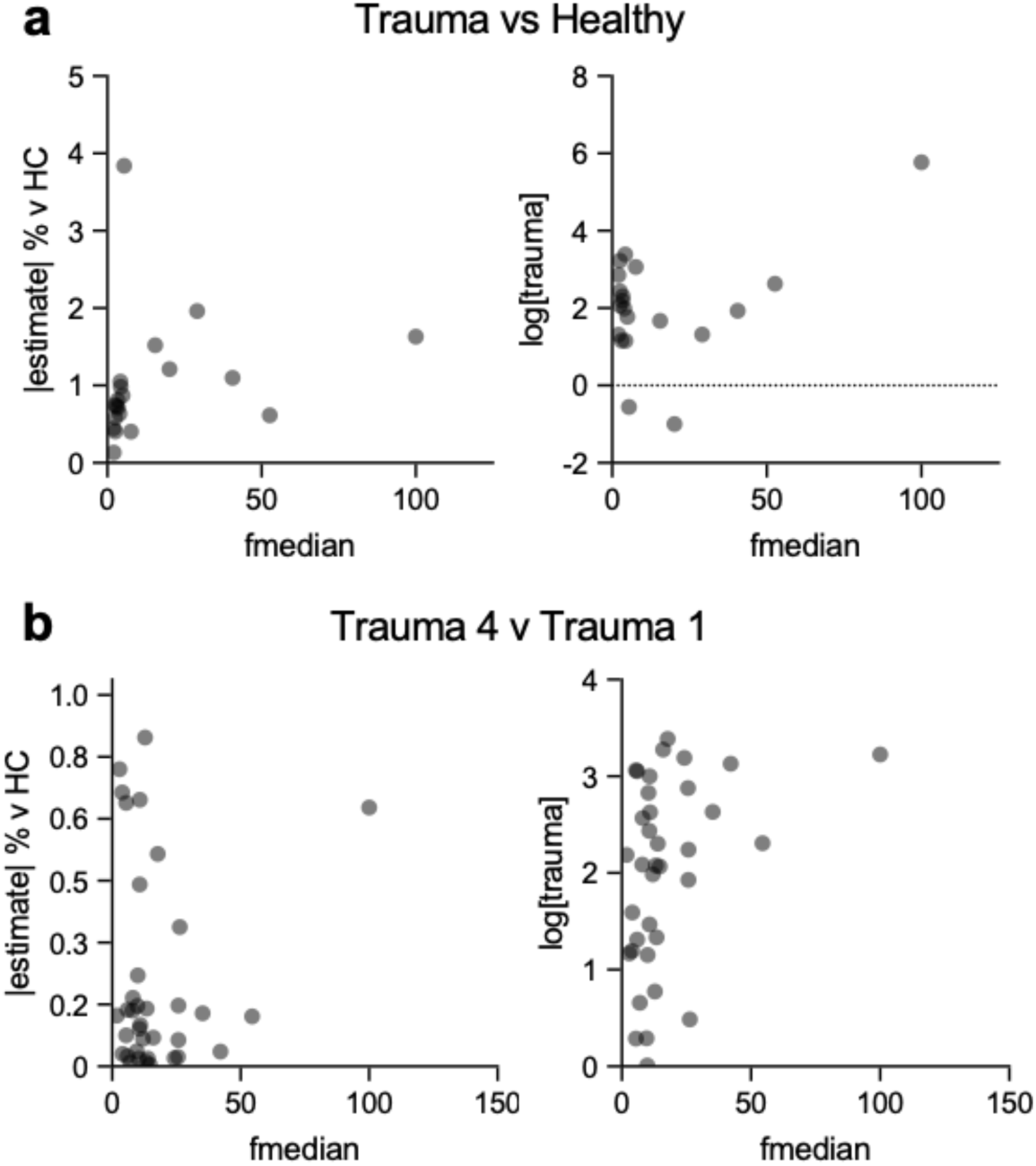
| Association of median relative importance with absolute percent change and analyte concentration. (a) Median relative importance (fmedian) of proteins across 10 model iterations versus its absolute percent change of concentration in trauma patients versus healthy controls (left) and concentration of analyte (right) in comparison between healthy controls and trauma. (b) Median relative importance (fmedian) of proteins across 10 model iterations versus its absolute percent change of concentration from trauma level 1 to trauma level 4 (left) and concentration of analyte (right) in comparison between trauma level.

**Supplemental Figure 11.**
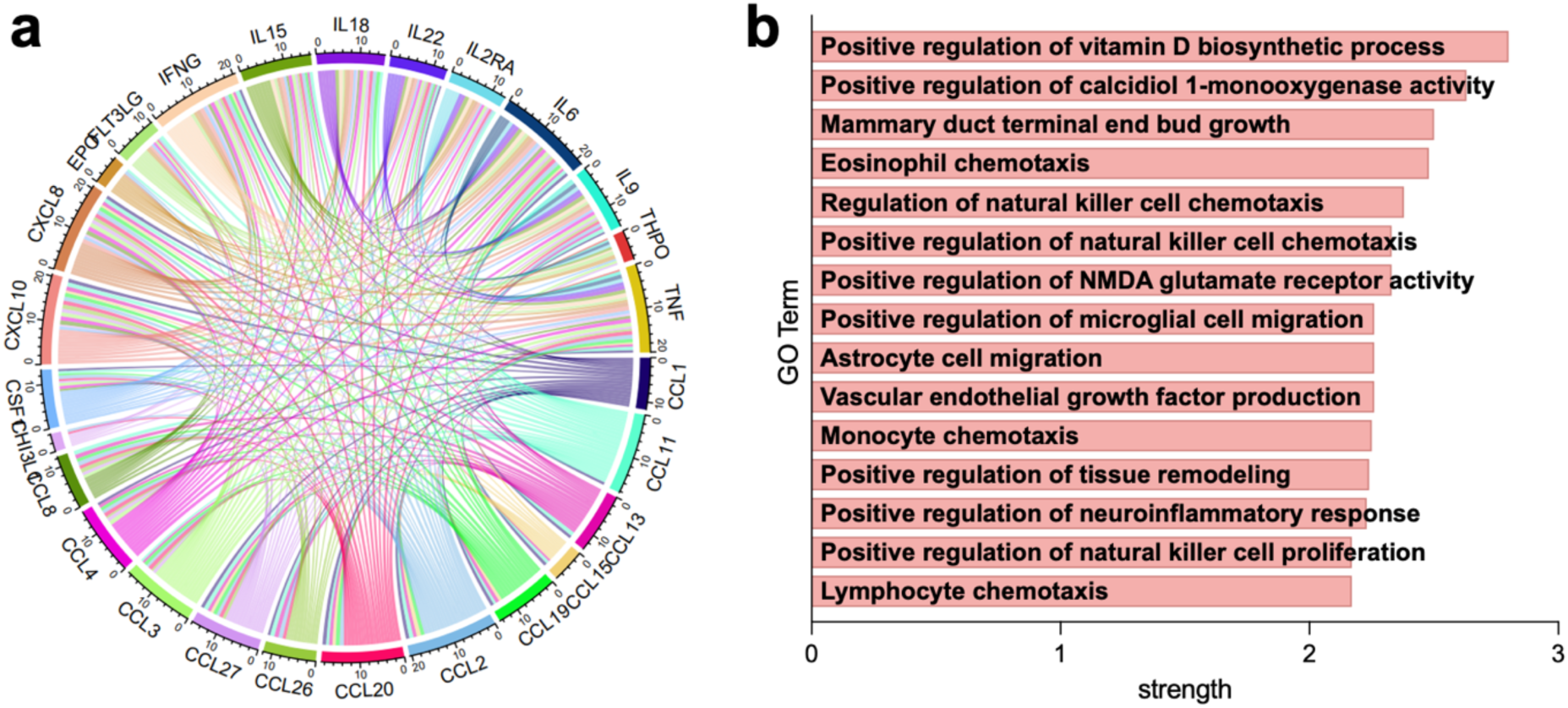
| STRING analysis of proteins upregulated with age. (a) Chord diagram generated based on STRING analysis of interactions among proteins upregulated with age. (b) STRING database gene ontology enrichment of biological processes from proteins upregulated with age (top 15 in strength of enrichment, FDR < 0. 05).

**Supplemental Figure 12.**
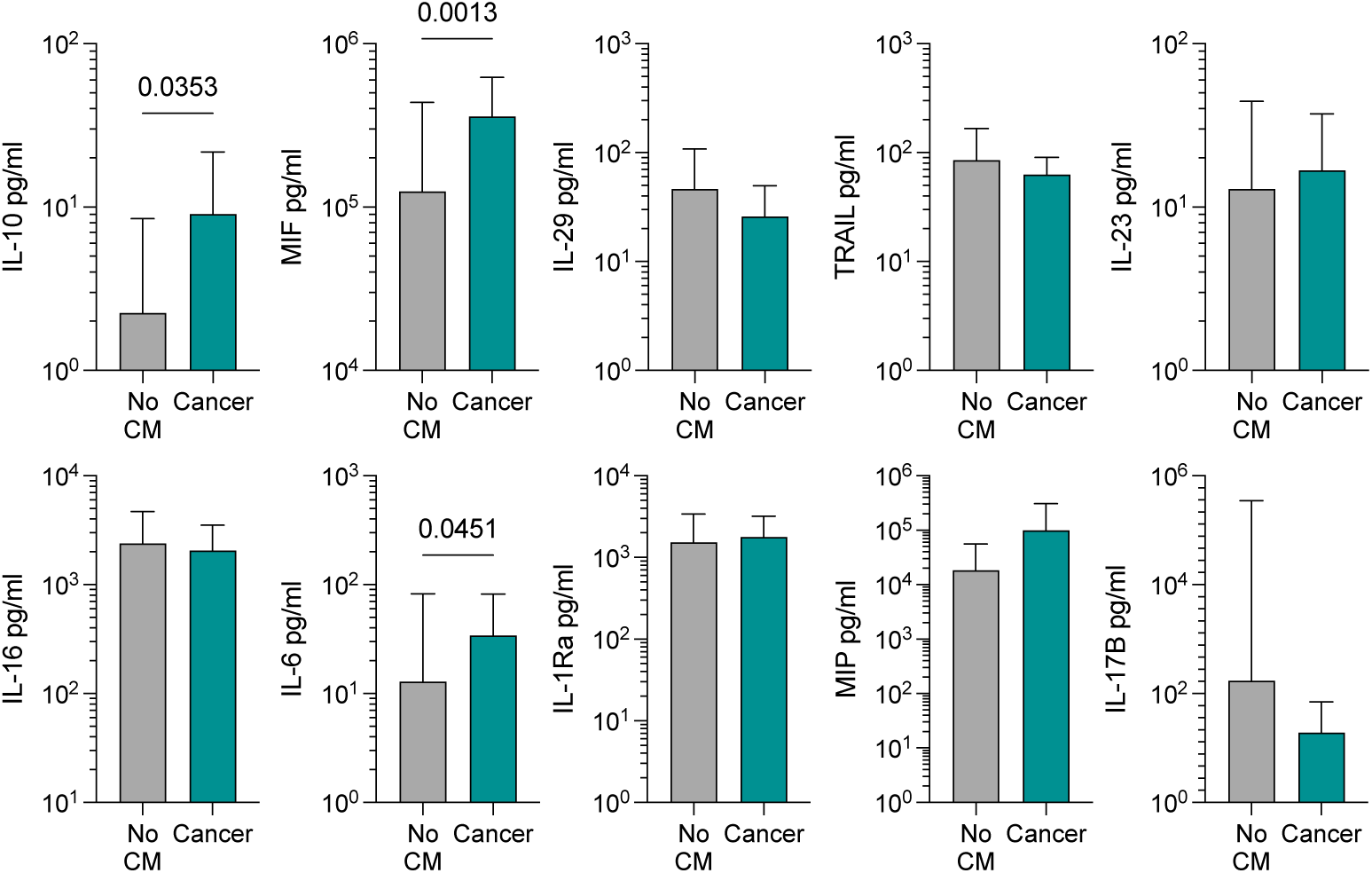
| Exploratory analyses of trauma patients with no comorbidities versus those with reported cancer incidence. Grey = no comorbidities (CM), Teal = cancer patients (cancer). Data are mean ± standard deviation, p = student’s T test (without correction for multiple comparisons).

**Supplemental Table 1.** | Demographics and basic trauma information for patients. Sex = biologic sex. Race = Self-reported race (EA = European American, AA = African American, AsA = Asian American). Ethnicity = Self-reported ethnicity (H/L = Hispanic/Latino). MVC = motor vehicle crash, GSW = gunshot wound or shotgun wound.

|  | Count | % |
| --- | --- | --- |
| **Location** |  |  |
| Worcester | 154 | 15.45 |
| Charlotte | 219 | 21.97 |
| Baltimore | 208 | 20.86 |
| Jacksonville | 223 | 22.37 |
| Miami | 193 | 19.36 |
| **Sex** |  |  |
| Male | 725 | 72.50 |
| Female | 272 | 27.20 |
| Unknown | 3 | 0.30 |
| **Race** |  |  |
| White or EA | 552 | 55.20 |
| Black or AA | 375 | 37.50 |
| Asian or AsA | 6 | 0.60 |
| Native American/Alaska Native | 6 | 0.60 |
| Other | 28 | 2.80 |
| Unknown | 33 | 3.30 |
| **Ethnicity** |  |  |
| Hispanic/Latino | 162 | 16.20 |
| Not H/L | 818 | 81.80 |
| Unknown | 20 | 2.00 |
| **Age** |  |  |
| 18 - 39 | 499 | 50.51 |
| 40 - 70 | 373 | 37.75 |
| > 70 | 113 | 11.44 |
| Unknown | 3 | 0.30 |
| **Mechanism of Injury** |  |  |
| MVC | 448 | 37.87 |
| Fall | 221 | 18.68 |
| GSW | 117 | 9.89 |
| Assault | 221 | 18.68 |
| Stab | 54 | 4.56 |
| Fire/burn | 17 | 1.44 |
| Nature/environment | 3 | 0.25 |
| Non-Motorized Transport | 4 | 0.34 |
| Drowning | 1 | 0.08 |
| Other Motorized Transport | 16 | 1.35 |
| Other, specified | 77 | 6.51 |
| Other, unspecified | 1 | 0.08 |
| Unknown | 3 | 0.25 |
| **Tested for COVID?** |  |  |
| Yes | 652 | 65.20 |
| No | 332 | 33.20 |
| Unknown/NA | 16 | 1.60 |

**Supplemental Table 2.**
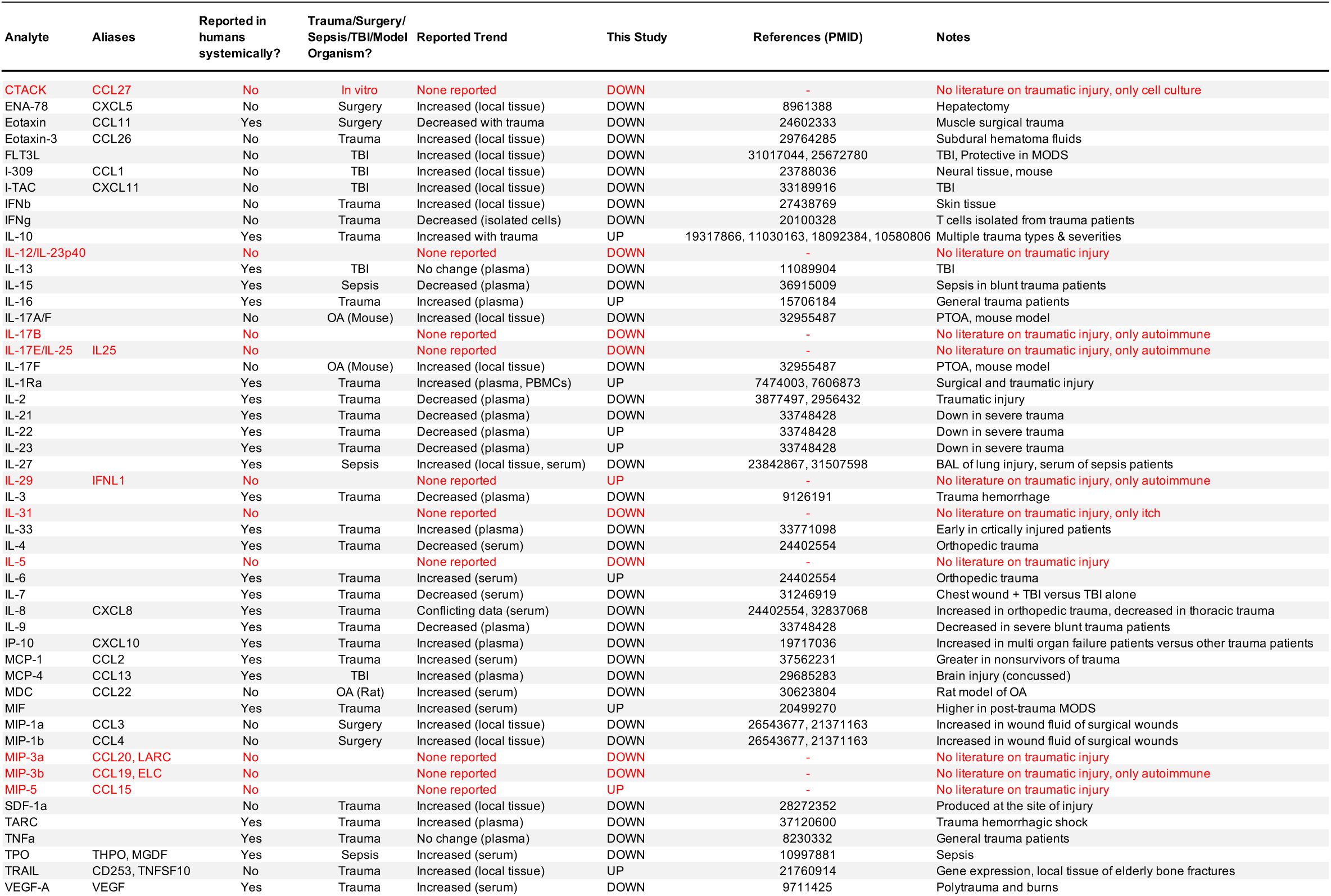
| Literature review of cytokines analyzed and their reported trends in trauma and associated conditions. Review on both GoogleScholar and PubMed search engines using key words “trauma”. “injury”, “wound”, “traumatic injury”, with or without “human”. Publications are only listed that are indexed in PubMed.

**Supplemental Table 3.** | Literature review of cytokines analyzed and their reported trends with age in human subjects. Review on both GoogleScholar and PubMed search engines using key words “age”, “increase”, “decrease”, “correlate with age”, and “human” or “patient”. Publications are only listed that are indexed in PubMed.

| Analyte | Aliases | Reported Trend with Age | This Study | References (PMID) | Notes |
| --- | --- | --- | --- | --- | --- |
| CTACK | CCL27 | None reported | UP | - |  |
| ENA-78 | CXCL5 | None reported | DOWN | - |  |
| EPO |  | Increase with age | UP | 25915923 |  |
| Eotaxin | CCL11 | Increase with age | UP | 26080062 |  |
| Eotaxin-3 | CCL26 | Decrease with age | UP | 30685456 | Dermatitis patients |
| FLT3L |  | Increase with age | UP | 26080062 |  |
| Gro-a | CXCL1 | None reported | DOWN | - |  |
| I-309 | CCL1 | None reported | UP | - |  |
| IFNg |  | Increase with age | UP |  | In children |
| IL-12/IL-23p40 |  | Increase with age | UP | 10671301 | Evaluated total IL-12 |
| IL-15 |  | Increase with age | UP | 16192677 |  |
| IL-16 |  | None reported | DOWN | - |  |
| IL-18 |  | Increase with age | UP | 21571262 | Secreted from dendritic cells (not total) |
| IL-22 |  | None reported | UP | - |  |
| IL-27 |  | Trended increase | UP |  |  |
| IL-2Ra | CD25 | None reported | UP | - |  |
| IL-6 |  | Debated | UP | 1453878, 11213271 | Some reports of increase, some decrease with age |
| IL-8 | CXCL8 | None reported | UP | - |  |
| IL-9 |  | None reported | UP | - |  |
| IP-10 | CXCL10 | Increase with age | UP | 26080062 | Trauma Patients |
| M-CSF | CSF1 | Trended increase | UP | 26080062 |  |
| MCP-1 | CCL2 | Increase with age | UP | 23039889 | Study incorrectly calls MCP-1 as CCL1, cited here under MCP-1 |
| MCP-2 | CCL8 | None reported | UP | - |  |
| MCP-4 | CCL13 | Increase with age | UP | 26080062 |  |
| MIP-1a | CCL3 | None reported | UP | - |  |
| MIP-1b | CCL4 | Decrease with age | UP | 30448299 | Trauma Patients |
| MIP-3a | CCL20, LARC | None reported | UP | - |  |
| MIP-3b | CCL19, ELC | None reported | UP | - |  |
| MIP-5 | CCL15 | None reported | UP | - |  |
| TNFa |  | Increase with age | UP | 10931139 |  |
| TPO | THPO, MGDF | None reported | UP | - |  |
| TRAIL | CD253, TNFSF10 | None reported | DOWN | - |  |
| VEGF-A | VEGF | Increase with age | UP | 26080062 |  |
| YKL-40 | CHI3L1 | Increase with age | UP | 18070151 |  |

**Supplemental Table 4.**
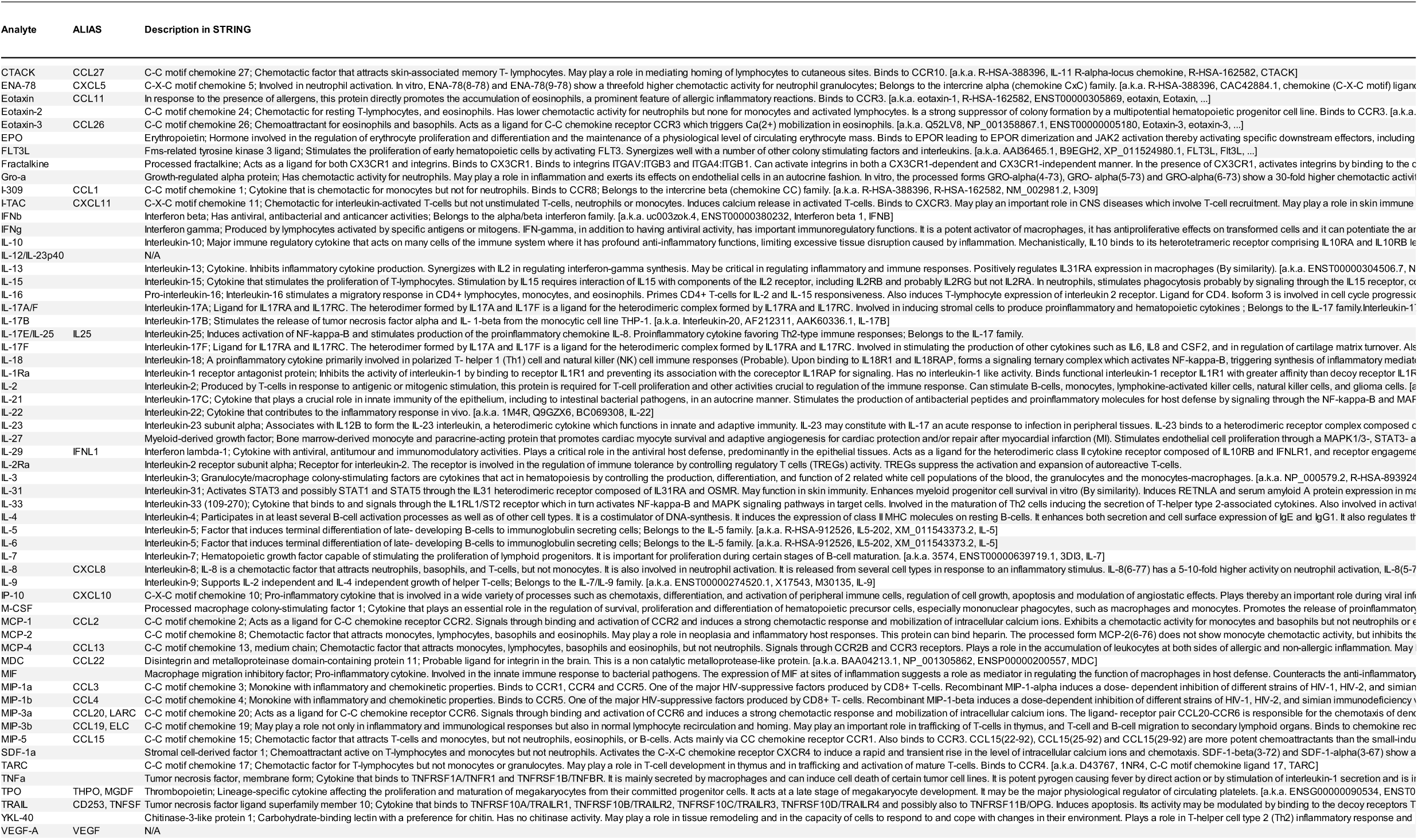
| STRING description of each protein. Protein descriptions from STRING database.

**Supplemental Table 5.** | Confusion Matrices for Models Applied to Determine Survival. Number of true negatives, false negatives, true positives, and false positives from a sample subset used to test the accuracy of machine learning models predicting death based on the expression levels of the 5-analyte panel.

|  | True<br>Negative | False<br>Positive | False<br>Negative | True<br>Positive |
| --- | --- | --- | --- | --- |
| Logistic Regression with Class Weights | 121 | 19 | 2 | 3 |
| Logistic Regression with SMOTE | 114 | 26 | 1 | 4 |
| Random Forest with Class Weights | 137 | 3 | 4 | 1 |
| Gradient Boosting with Class Weights | 138 | 2 | 2 | 3 |
| XGBoost with Class Weights | 132 | 8 | 2 | 3 |

## SUPPLEMENTAL METHODS

### Random Forest Model Tuning

Data was split 70/30 into training/test datasets, maintaining the ratio of trauma:healthy patients in each set. Hyperparameters were tuned with tidymodels grid search (n=20) to determine the appropriate number of variables to randomly sample at splitting (mtry) and minimum amount of data needed to split (min_n). Tuning was done with 10 trees using 5-fold cross validation to select the model with the highest AUC-ROC as the best model. This model was trained on the training set, then applied to test data. It was then validated on both training and test data separately with the same tuned mtry and min_n parameters and 200 trees. Predictions, model metrics, and variable permutation importance values from these final models were collected.

### Random Forest Variable Importance

Permutation importance calculates the influence of each variable on model prediction based on how much a change in the variable’s values affects the model’s predictive error.^43^ To derive summary statistics of variable importance values and to minimize the chance of a variable erroneously showing up as important, the model training and validation steps were repeated on the dataset 10 times from different seeds and the permutation importance for each variable was ranked for each iteration. The raw variable importance values were then converted to relative importance by dividing each variable importance score by the largest importance score of the variables for each of the 10 iterations and multiplied by 100.^54^ Variables whose permutation importance ranked in the top 10 in at least 5 of the iterations were plotted, along with their minimum, first quartile, median, mean, third quartile, and maximum importance values.

### Confirmation of literature findings

Our observation of decreased IL-2 levels, with many below the LOD, in trauma patients versus control samples is consistent with reports of reduced IL-2 production (associated with T cell activation) in response to traumatic injury.^64,65^ In agreement with the literature, trauma patients also exhibited increased levels of several other interleukins, including IL-16,^66,67^ IL-1Ra,^68,69^ and IL-6,^3^ as well as a decrease in IL-21^4^ and IL-3.^70^ We observed several other patterns consistent with previous literature. These include MIF (upregulated^71^), TRAIL (upregulated and previously reported to be increased in elderly osteoporotic fractures^72^), and Eotaxin (downregulated, and previously reported to be downregulated in musculoskeletal surgical trauma^67,73^).

### Downregulated proteins that are contrary to literature findings

We also observed some trends that differ from those reported in the literature. The downregulation of VEGF-A, which is involved in angiogenesis, in trauma samples compared to healthy samples is especially interesting due to the involvement of angiogenesis in tissue repair. Although previous studies have demonstrated increased serum VEGF levels on day of arrival for both trauma and burn patients,^74^ we observed a lower level of VEGF-A in trauma samples, regardless of injury mechanism, general wound type, specific wound type, wound location, or trauma level. This suggests a potential decrease in angiogenesis in the immediate aftermath of injury, possibly as a mechanism to control blood loss.

Of the other proteins for which we found trends that are contrary to what was previously published in the literature, evaluation of many of these proteins in human patients had primarily been limited to TBI. Increased levels of FLT3L, which activates FLT3 to stimulate proliferation of hematopoietic cells, have been reported in a study of the cerebral fluid of 10 patients that had experienced severe TBI.^20^ In mice, FLT3L has been shown to exhibit protective effects in ischemia-reperfusion injury, volumetric muscle loss, and multi-organ dysfunction syndrome.^75–77^ We observed significant decreases in FLT3L levels in trauma, as well as for each injury mechanism, general wound type, specific wound type, and wound location, versus healthy controls. This was also true for patients who had head/neck wounds, although it cannot be assumed that those patients would have TBI. Thus, the type of injury (ex. TBI vs. Non-TBI) likely played a role in the differences in our observation compared to the previous reports. In a study of brain tissue samples from 12 patients with severe TBI, I-TAC, which is chemotactic for IL-activated T-cells, was found to be upregulated. Although we observed an upregulation of I-TAC in trauma patients compared to healthy controls, this difference was not statistically significant. Nevertheless, other factors such as the use of systemic fluids (I.e. blood) versus localized fluids or tissues should also be taken into consideration, as it is possible that local elevation of proteins would not be reflected in systemically-sampled fluid.

Levels of IL-7, which is involved in stimulation of lymphoid progenitors, have previously been reported to be elevated with increasing injury severity for trauma patients,^4^ although we observed a significantly lower level of IL-7 for trauma patients versus controls, and no significant difference between IL-7 levels in the most severe (level 1) cases compared to less severe (levels 2, 3, or 4) cases. However, this is likely a result of our inclusion of non-ICU patients, while the Cai et al. study included only trauma patients who were admitted to the ICU and survived to discharge. Another study found that TBI patients with chest injury had lower mean IL-7 levels than TBI patients without chest injury.^19^ Although we observed a significant decrease in IL-7 levels across all wound types, wound locations, and injury mechanisms, these findings altogether suggest the importance of injury characteristics on IL-7 levels. This is further supported by conflicting reports on the levels of a number of other cytokines, which is likely a result of differences in patient characteristics. For example, IL-8 levels were shown to be higher in patients with orthopedic injures compared to healthy controls,^3^ while serum IL-8 levels were shown to be higher in healthy controls compared to chest trauma patients.^78^ Meanwhile, we observed lower IL-8 levels for trauma patients across all injury mechanisms, wound types, and wound locations, compared to healthy controls. TPO also has conflicting reports in the literature, with one study finding that patients with multiple trauma have elevated serum TPO in the days following injury,^79^ while another study found that TPO levels were not measurable for patients with major lower extremity trauma.^80^ Lower TPO levels have also been associated with nonsurvival in patients that have experienced major trauma.^81^ We observed significantly lower TPO in trauma patients compared to healthy controls, regardless of injury mechanism, wound type, and wound location, However, patients with the most severe injuries exhibited higher TPO levels than patients with the least severe injuries, which appears to support prior findings of elevated TPO after severe injury.

A statistically significant decrease in concentration of IL-17E/IL-25 was observed for injury mechanisms of GSW and MVC but was not statistically significant for the other injury mechanisms. Additionally, the wound type played a role in the concentration of IL-17E/IL-25. Patients who experienced only internal wounds or only penetrating wounds did not exhibit a significant decrease in this protein, but those who experienced both wound types did. We observed that reduced IL-17E/IL-25 concentration was statistically significant for patients who experienced central wounds or combined head/neck and central wounds, but not for patients who experienced combined head/neck and peripheral wounds, head/neck wounds only, combined peripheral and central wounds, or peripheral wounds only.

### Novel reporting of proteins with changes in levels below the LOD

The concentration of several proteins (IL-31, IL-5, MIP-3a) previously unreported in this setting were not significantly associated with trauma; however, there were significant differences in our assay’s ability to detect a signal. Each of these proteins were significantly less likely to be detected in trauma samples compared to healthy controls. This suggests an exceptional potential reduction in the concentrations of these proteins. IL-31 is linked both to Th2 immunity associated with wound healing and to dermatitis. IL-5 is also associated with type-2 immunity and eosinophil inflammation. MIP-3a has been associated with autoimmunity in mouse models, and also downregulated with IL-10 which correlates with trauma that induces IL-10. Type-2 immune proteins have important impact on wound closure and collagen deposition, which are critical in long-term recovery of injuries. Disruption of these pathways presents another mechanism by which severe trauma limits healing ability by decreasing the induction of collagen stimulating immune responses to close wounds and strength those closures. Induction of these responses locally may provide a therapeutic window to assist in wound healing that may be compromised in patients sustaining severe trauma.

### Upregulated proteins that are contrary to literature findings

While levels of both IL-22 and IL-23 have been reported to decrease in severe trauma,^4^ we did not observe a significant change in IL-22 levels between trauma and healthy control patients. However, we found a significant increase in IL-23 concentration of trauma samples compared to healthy controls, which held true for patients injured through falls or stabbings, but not those who were injured in MVC or GSW. IL-23 levels are significantly higher in more severe (level 1) injuries, compared to less severe (level 3, level 4) injuries, and IL-22 levels were also significantly higher in level 1 versus level 4 injuries. Increased IL-23 levels were observed for patients who experienced only internal soft tissue wounds, or only penetrating soft tissue wounds, but not for patients with bone involvement. The observed differences between trends of IL-22 and IL-23 levels in our study compared to the previous report may be attributed to differences in patient cohorts, as the Cai et al. cohorts were comprised of only patients admitted to the ICU and who survived to discharge.

## REFERENCES

1. Sousa, A. et al. Measurement of Cytokines and Adhesion Molecules in the First 72 Hours after Severe Trauma: Association with Severity and Outcome. Disease Markers 2015, 1–8 (2015).

2. Neidhardt, R. et al. Relationship of Interleukin-10 Plasma Levels to Severity of Injury and Clinical Outcome in Injured Patients: *The Journal of Trauma: Injury*, Infection, and Critical Care 42, 863–871 (1997).

3. Volpin, G. et al. Cytokine Levels (IL-4, IL-6, IL-8 and TGFβ) as Potential Biomarkers of Systemic Inflammatory Response in Trauma Patients. International Orthopaedics (SICOT) 38, 1303–1309 (2014).

4. Cai, J., et al. Protective/reparative cytokines are suppressed at high injury severity in human trauma. Trauma Surg Acute Care Open 6, e000619 (2021).

5. Lord, J. M. et al. The systemic immune response to trauma: an overview of pathophysiology and treatment. The Lancet 384, 1455–1465 (2014).

6. Sadtler, K., Collins, J., Byrne, J. D. & Langer, R. Parallel evolution of polymer chemistry and immunology: Integrating mechanistic biology with materials design. Advanced Drug Delivery Reviews 156, 65–79 (2020).

7. Ngo, T. B. et al. SARS-CoV-2 Seroprevalence and Drug Use in Trauma Patients from Six Sites in the United States. Preprint at 10.1101/2021.08.10.21261849 (2021).

8. Thomas, F. D., et al. Drug and Alcohol Prevalence in Seriously and Fatally Injured Road Users Before and During the COVID-19 Public Health Emergency. (2020).

9. F. D. Thomas et al. Alcohol and Drug Prevalence Among Seriously or Fatally Injured Road Users. (2022).

10. DO Ricke & C Aguilar. Multiscale analysis of a regenerative therapy for treatment of volumetric muscle loss injury. Gene Expression Omnibus (2018).

11. PV.018_L_PT2 RNA-Seq. Gene Expression Omnibus (2018).

12. DO Ricke & C Aguilar. In vivo Monitoring of Transcriptional Dynamics After Lower-Limb Muscle Injury Enables Quantitative Classification of Healing. Gene Expression Omnibus (2017).

13. Tanya J Shaw. Mouse 1 Back Wound [M1BW]. Gene Expression Omnibus (2020).

14. Aguilar, C. A. et al. Multiscale analysis of a regenerative therapy for treatment of volumetric muscle loss injury. Cell Death Discovery 4, 33 (2018).

15. Greising, S. M. et al. Unwavering Pathobiology of Volumetric Muscle Loss Injury. Sci Rep 7, 13179 (2017).

16. Aguilar, C. A. et al. In vivo Monitoring of Transcriptional Dynamics After Lower-Limb Muscle Injury Enables Quantitative Classification of Healing. Sci Rep 5, 13885 (2015).

17. Usansky, I. et al. A developmental basis for the anatomical diversity of dermis in homeostasis and wound repair. The Journal of Pathology 253, 315–325 (2021).

18. Johansen, J. S. et al. High serum YKL-40 level in a cohort of octogenarians is associated with increased risk of all-cause mortality. Clinical and Experimental Immunology 151, 260– 266 (2008).

19. Crawford, A. M. et al. Concomitant chest trauma and traumatic brain injury, biomarkers correlate with worse outcomes. J Trauma Acute Care Surg 87, S146–S151 (2019).

20. Dyhrfort, P. et al. Monitoring of Protein Biomarkers of Inflammation in Human Traumatic Brain Injury Using Microdialysis and Proximity Extension Assay Technology in Neurointensive Care. Journal of Neurotrauma 36, 2872–2885 (2019).

21. Mousessian, A. S. et al. CXCR7, CXCR4, and Their Ligand Expression Profile in Traumatic Brain Injury. World Neurosurgery 147, e16–e24 (2021).

22. Lok Ting Lau & Albert Cheung-Hoi Yu. Astrocytes Produce and Release Interleukin-1, Interleukin-6, Tumor Necrosis Factor Alpha and Interferon-Gamma Following Traumatic and Metabolic Injury. Journal of Neurotrauma 18, (2004).

23. Meabon, J. S. et al. Chronic elevation of plasma vascular endothelial growth factor-A (VEGF-A) is associated with a history of blast exposure. Journal of the Neurological Sciences 417, 117049 (2020).

24. Timmermans, K. et al. Plasma levels of danger-associated molecular patterns are associated with immune suppression in trauma patients. Intensive Care Med 42, 551–561 (2016).

25. Huangfu, L., Li, R., Huang, Y. & Wang, S. The IL-17 family in diseases: from bench to bedside. Sig Transduct Target Ther 8, 402 (2023).

26. Riedel, J.-H. et al. IL-17F Promotes Tissue Injury in Autoimmune Kidney Diseases. JASN 27, 3666–3677 (2016).

27. Majumder, S. & McGeachy, M. J. IL-17 in the Pathogenesis of Disease: Good Intentions Gone Awry. Annu. Rev. Immunol. 39, 537–556 (2021).

28. Colletti, L. M., Kunkel, S. L., Green, M., Burdick, M. & Strieter, R. M. HEPATIC INFLAMMATION FOLLOWING 70% HEPATECTOMY MAY BE RELATED TO UP-REGULATION OF EPITHELIAL NEUTROPHIL ACTIVATING PROTEIN-78. Shock 6, (1996).

29. Colletti, L. M. et al. Chemokine expression during hepatic ischemia/reperfusion-induced lung injury in the rat. The role of epithelial neutrophil activating protein. J. Clin. Invest. 95, 134– 141 (1995).

30. Cooper, A. M. & Khader, S. A. IL-12p40: an inherently agonistic cytokine. Trends in Immunology 28, 33–38 (2007).

31. Shivaprasad, B. M. & Pradeep, A. R. Effect of non-surgical periodontal therapy on interleukin-29 levels in gingival crevicular fluid of chronic periodontitis and aggressive periodontitis patients. Disease Markers 34, (2013).

32. Saif Salahuddin Jasim & Ghada Ibrahim Taha. Role of type III Interferon (IL-29) on HSV-1 infection in breast cancer patients suffering from periodontitis and receiving chemotherapy. ijbd 16, 85–95 (2023).

33. Lotfi, W., et al. Serum level of Interleukin 29 in verruca vulgaris: Case control study. Fayoum University Medical Journal 3, 32–35 (2019).

34. Aya Refat, E. & Zainab Hussein, A.-H. IL-24 AND IL-29 IN T2DM WITH AND WITHOUT DIABETIC FOOT ULCERS. achi 30, 103–119 (2022).

35. Gilliver, S. C., Ruckshanthi, J. P. D., Hardman, M. J., Nakayama, T. & Ashcroft, G. S. Sex Dimorphism in Wound Healing: The Roles of Sex Steroids and Macrophage Migration Inhibitory Factor. Endocrinology 149, 5747–5757 (2008).

36. Kruse, J. L. et al. Inflammation and depression treatment response to electroconvulsive therapy: Sex-specific role of interleukin-8. *Brain*, Behavior, and Immunity 89, 59–66 (2020).

37. Kruse, J. L. et al. Depression treatment response to ketamine: sex-specific role of interleukin-8, but not other inflammatory markers. Transl Psychiatry 11, 167 (2021).

38. Da Pozzo, E., Giacomelli, C., Cavallini, C. & Martini, C. Cytokine secretion responsiveness of lymphomonocytes following cortisol cell exposure: Sex differences. PLoS ONE 13, e0200924 (2018).

39. Kim, D. H. J. et al. Neonatal immune signatures differ by sex regardless of neurodevelopmental disorder status: Macrophage migration inhibitory factor (MIF) alone reveals a sex by diagnosis interaction effect. Brain, Behavior, and Immunity 111, 328–333 (2023).

40. Larsson, A. et al. The effects of age and gender on plasma levels of 63 cytokines. Journal of Immunological Methods 425, 58–61 (2015).

41. Zhou, L. et al. Age-specific changes in the molecular phenotype of patients with moderate-to-severe atopic dermatitis. Journal of Allergy and Clinical Immunology 144, 144–156 (2019).

42. Decker, M.-L., Gotta, V., Wellmann, S. & Ritz, N. Cytokine profiling in healthy children shows association of age with cytokine concentrations. Sci Rep 7, 17842 (2017).

43. Rea, I. M., McNerlan, S. E. & Alexander, H. D. TOTAL SERUM IL-12 AND IL-12p40, BUT NOT IL-12p70, ARE INCREASED IN THE SERUM OF OLDER SUBJECTS; RELATIONSHIP TO CD3+AND NK SUBSETS. Cytokine 12, 156–159 (2000).

44. Gangemi, S. et al. Age-Related Modifications in Circulating IL-15 Levels in Humans. Mediators of Inflammation 2005, 245–247 (2005).

45. Ciaramella, A. et al. Effect of age on surface molecules and cytokine expression in human dendritic cells. Cellular Immunology 269, 82–89 (2011).

46. Lamparello, A. J., Namas, R. A., Abdul-Malak, O., Vodovotz, Y. & Billiar, T. R. Young and Aged Blunt Trauma Patients Display Major Differences in Circulating Inflammatory Mediator Profiles after Severe Injury. Journal of the American College of Surgeons 228, 148–160e7 (2019).

47. Mansfield, A. S. et al. Normal ageing is associated with an increase in Th2 cells, MCP-1 (CCL1) and RANTES (CCL5), with differences in sCD40L and PDGF-AA between sexes. Clinical and Experimental Immunology 170, 186–193 (2012).

48. Bruunsgaard, H., SKINHéJ, P., Pedersen, A. N., Schroll, M. & Pedersen, B. K. Ageing, tumour necrosis factor-alpha (TNF-a) and atherosclerosis. Clinical and Experimental Immunology (2000).

49. Ishiguro, A. et al. Age-related changes in thrombopoietin in children: reference interval for serum thrombopoietin levels. Br J Haematol 106, 884–888 (1999).

50. Shin, M. S. et al. Maintenance of CMV-specific CD8+ T cell responses and the relationship of IL-27 to IFN-γ levels with aging. Cytokine 61, 485–490 (2013).

51. Niwa, Y., Kasama, T., Miyachi, Y. & Kanoh, T. NEUTROPHIL CHRMOTAXIS, PHAGOCYTOSIS AND PARAMETERS OF REACTIVE OXYGEN SPECIES IN HUMAN AGING: CROSS-SECTIONAL AND LONGITUDINAL STUDIES. 44, (1989).

52. Wenisch, C., Patruta, S., Daxböck, F., Krause, R. & Hörl, W. Effect of age on human neutrophil function. Journal of Leukocyte Biology 67, 40–45 (2000).

53. Martínez De Toda, I., Maté, I., Vida, C., Cruces, J. & De La Fuente, M. Immune function parameters as markers of biological age and predictors of longevity. Aging 8, 3110–3119 (2016).

54. Namas, R. A. et al. Temporal Patterns of Circulating Inflammation Biomarker Networks Differentiate Susceptibility to Nosocomial Infection Following Blunt Trauma in Humans. Annals of Surgery 263, 191–198 (2016).

55. Baucom, M. R. et al. Predictive Value of Early Inflammatory Markers in Trauma Patients Based on Transfusion Status. Journal of Surgical Research 291, 691–699 (2023).

56. Rallis, K. S. et al. IL-10 in cancer: an essential thermostatic regulator between homeostatic immunity and inflammation – a comprehensive review. Future Oncology 18, 3349–3365 (2022).

57. Szklarczyk, D. et al. The STRING database in 2023: protein–protein association networks and functional enrichment analyses for any sequenced genome of interest. Nucleic Acids Research 51, D638–D646 (2023).

58. Kleiber, C. & Zeileis, A. AER: Applied Econometrics with R. (2024).

59. Storey, J. D. The positive false discovery rate: a Bayesian interpretation and the q-value. Ann. Statist. 31, (2003).

60. Krijthe, J. H. Rtsne: T-Distributed Stochastic Neighbor Embedding using a Barnes-Hut Implementation. (2015).

61. Kuhn, M. & Wickham, H. Tidymodels: a collection of packages for modeling and machine learning using tidyverse principles.

62. Mayer, L. M. et al. Machine Learning in Infectious Disease for Risk Factor Identification and Hypothesis Generation: Proof of Concept Using Invasive Candidiasis. Open Forum Infectious Diseases 9, ofac401 (2022).

63. R Core Team. The R Project for Statistical Computing. (2022).

64. Faist, E. et al. Prostaglandin E2 (PGE2)-dependent Suppression of Interleukin α (IL-2) Production in Patients with Major Trauma. The Journal of Trauma and Acute Care Surgery 27, (1987).

65. Edward, A. & Regan, R. F. The Effects of Hemorrhage and Trauma on Interleukin 2 Production. Arch Surg 120, (1985).

66. Shimonkevitz, R., Northrop, J., Harris, L., Craun, M. & Bar-Or, D. Interleukin-16 Expression in the Peripheral Blood and CD8 T Lymphocytes After Traumatic Injury: *The Journal of Trauma: Injury*, Infection, and Critical Care 58, 252–258 (2005).

67. Reikeras, O., Borgen, P., Reseland, J. E. & Lyngstadaas, S. P. Changes in serum cytokines in response to musculoskeletal surgical trauma. BMC Res Notes 7, 128 (2014).

68. Ertel, W. et al. Release of Anti-inflammatory Mediators after Mechanical Trauma Correlates with Severity of Injury and Clinical Outcome: *The Journal of Trauma: Injury*, Infection, and Critical Care 39, 879–887 (1995).

69. Nualláin, E. M. Ó., Puri, P., Mealy, K. & Reen, D. J. Induction of Interleukin-1 Receptor Antagonist (IL-1ra) Following Surgery Is Associated with Major Trauma. Clinical Immunology and Immunopathology 76, (1995).

70. Xu, Y. X., Wichmann, M. W., Ayala, A., Cioffi, W. G. & Chaudry, I. H. Trauma–Hemorrhage Induces Increased Thymic Apoptosis While Decreasing IL-3 Release and Increasing GM-CSF. Journal of Surgical Research 68, 24–30 (1997).

71. Joshi, P. C., Poole, G. V., Sachdev, V., Zhou, X. & Jones, Q. Trauma patients with positive cultures have higher levels of circulating macrophage migration inhibitory factor (MIF). Res Commun Mol Pathol Pharmacol. 107, (2000).

72. Zhang, Y.-P. et al. Pathway-Based Association Analyses Identified TRAIL Pathway for Osteoporotic Fractures. PLoS ONE 6, e21835 (2011).

73. Bastian, D., Tamburstuen, M. V., Lyngstadaas, S. P. & Reikerås, O. Local and systemic chemokine patterns in a human musculoskeletal trauma model. Inflamm. Res. 58, 483–489 (2009).

74. Grad, S. et al. Strongly Enhanced Serum Levels of Vascular Endothelial Growth Factor (VEGF) after Poly-trauma and Burn. cclm 36, 379–383 (1998).

75. Tian, G., Lu, J., Guo, H., Liu, Q. & Wang, H. Protective effect of Flt3L on organ structure during advanced multiorgan dysfunction syndrome in mice. Molecular Medicine Reports 11, 4135–4141 (2015).

76. Zhou, C. et al. FLT3/FLT3L-mediated CD103+ dendritic cells alleviates hepatic ischemia-reperfusion injury in mice via activation of treg cells. Biomedicine & Pharmacotherapy 118, 109031 (2019).

77. Lokwani, R. et al. Pro-regenerative biomaterials recruit immunoregulatory dendritic cells after traumatic injury. Nat. Mater. 23, 147–157 (2024).

78. Bagaria, V. et al. Predicting Outcomes After Blunt Chest Trauma—Utility of Thoracic Trauma Severity Score, Cytokines (IL-1β, IL-6, IL-8, IL-10, and TNF-α), and Biomarkers (vWF and CC-16). Indian J Surg 83, 113–119 (2021).

79. Hobisch-Hagen, P. et al. Low platelet count and elevated serum thrombopoietin after severe trauma. European J of Haematology 64, 157–163 (2000).

80. Eric I. Choe et al. Thrombocytosis After Major Lower Extremity Trauma: Mechanism and Possible Role in Free Flap Failure. Ann Plast Surg 36, 489–494.

81. Schofield, H. et al. Immature platelet dynamics are associated with clinical outcomes after major trauma. Journal of Thrombosis and Haemostasis S1538783623008656 (2023) doi:10.1016/j.jtha.2023.12.002.

